# Exploring a digital health solution to collect and manage health-related needs for complex surgery patients: Mixed-methods study

**DOI:** 10.1101/2025.05.23.25328074

**Authors:** Naleef Fareed, Humza Asgher, Diamantis Tsilimigras, Odysseas P. Chatzipanagiotou, Giovanni Catalano, Bridget Hartwell, Kathleen Bolton, Timothy M. Pawlik

**Affiliations:** Department of Biomedical Informatics, College of Medicine, The Ohio State University, Columbus, Ohio, USA; Department of Surgery, College of Medicine, The Ohio State University, Columbus, Ohio, USA; The James Surgical Oncology Department, The Ohio State University, Columbus, Ohio, USA

**Keywords:** health-related needs, collection of social needs, surgery, digital health

## Abstract

**Background:** Patients who undergo complex surgery often experience substantial burden of health-related needs (medical, social, and behavioral health). A closed loop digital solution could facilitate the collection and resolution of health-related needs by care team members for patients who undergo complex surgery. A digital solution may facilitate adherence to a clear treatment plan and concomitantly reduce surgical complications and readmissions associated with unmet health-related needs, which remain persistent challenges across health care settings.

**Objectives:** To establish a set of user specifications for a digital solution to collect and manage health-related needs, specifically medical, social, and behavioral needs for patients who undergo complex surgery.

**Methods:** We applied the Double Diamond Framework and organized the study into two phases: (1) qualitative methods to discover patients’ and care team members’ perspectives on health-related needs; (2) participatory design sessions to gain feedback about ideal features of a digital solution. We supplemented both phases with analysis of electronic health record data.

**Results:** Extensive themes emerged from interviews with patients (n=20) and care team members (n=24), capturing their health-related and surgical experiences as well as desired features for a proposed digital solution. A subset of participants from Phase 1 (n=5 patients and n=9 care team members) provided feedback on preferred features, drawing from digital tools currently available in the electronic health record system at our academic medical center. Findings from the qualitative interviews and design sessions were corroborated with documentation in the electronic health record.

**Conclusion:** Digital solutions could provide a streamlined approach for collection and management of health-related needs in surgery, with the goal of addressing unmet needs and improving patient activation. This approach is critical to ensure patients, especially patients who undergo complex surgery, have positive health outcomes. We identified preferences for specific features in a proposed digital solution based on our systematic assessment that will inform future work.

## Introduction

The evidence on how to best collect and manage health-related needs is inadequate. Health care organizations in the United States are especially face challenges in implementing policies that increase requirements in reporting health-related needs.^1–4^ Furthermore, the integration of health-related social needs with medical and behavioral needs represents a system level effort to recognize the importance of whole-person health as a mechanism for transforming health care quality.^5,6^ The collection, integration, and management of health-related needs (i.e., medical, social, and behavioral) can be complicated and time intensive, especially with social needs requiring substantial resources to resolve unmet needs.^7^ Screening rates and documentation of social needs have greatly varied across clinics, and hospital systems require effective collaboration with referral organizations, such as community-based organizations to effectively address these needs.^8^

Studies on health-related needs, particularly social needs, have been conducted in internal medicine, pediatrics, and outpatient settings;^9,10^ however, effective screening in surgical care remains poorly defined. Social needs have been linked to complications and mortality during and soon after a surgical episode.^11^ Patients with unaddressed social needs are twice as likely to seek care in the emergency department,^12^ which may disrupt treatment goals, negatively affect recovery from surgery, and contribute to readmissions. Despite this, integration of medical, social, and behavioral needs to provide holistic care remains a significant challenge in surgery, like many other clinical settings, and data from real world implementation are limited.^13,14^

Digital solutions provide a promising and comprehensive approach to improve collection of health-related needs, particularly social needs. Digital solutions can collect real-time data^15^ and enable real-time management of needs that can enhance the surgical experience.^16^ Timely management of needs with digital technologies can help patients recover earlier post-surgery and shorten their hospital stay.^16^ Digital solutions allow personalized content for each patient^17^ which can further support an individualized approach to address specific patient needs. Novel solutions have been proposed in the literature to collect and refer patients to local resources using various digital health platforms.^18,19^

To enhance the collection and management of health-related needs, it is essential to understand the perspectives of both patients and care team members. The perspectives of these key stakeholders can help identify barriers and facilitators, ultimately refining interventions to promote acceptance of such tools and optimize both treatment goal management and health-related needs that can impact the goals. Extant literature indicates that screening of health-related needs, particularly social needs, is less prevalent in specialist visits compared with primary care settings as screening for these factors may be challenging to integrate into specialized workflows.^20,21^ Thus, implementing a standardized approach, which may be grounded in a technological solution, may facilitate screening across different specialties. To bridge the gap in the literature, we conducted a mixed methods study on both care team members and patients to assess convergent and divergent themes focused on health-related needs screening and management in surgical settings at our Academic Medical Center (AMC). The objective of this study was to identify feature specifications for a digital solution that can transform the existing workflow at our AMC to improve management of patients who undergo complex surgery. Specifically, we sought to: (1) establish problems and gaps in the collection, integration, and management of health-related needs in the current treatment approach for complex surgery patients at our AMC and (2) define desired specifications for a digital solution, informed by feedback referencing digital tools currently available in the existing electronic health record system.

## Methods

### The Double Diamond framework

In the current study, we followed the “Double Diamond” framework for our user-centered design approach.^22^ The Double Diamond framework can be applied to design and develop a digital solution based on formative evidence. The framework allowed us to define problems identified by care team members and patients in the collection and management of health-related needs, suggest digital solutions for participants, and gather user perceptions regarding design and usability. We explained our diagnose, discover, define, develop, and deliver phases below.

### Diagnose

Our research team first performed a scoping review of the literature for major gaps related to the collection and management of health-related needs in surgery, with a particular focus on social needs. Our study focused on experiences with health-related needs and preferences for digital solutions to report such needs. We reviewed various peer-reviewed articles to identify gaps in the current literature, including formative work for this study.^23^ We also reviewed our AMC EHR progress notes for patients who underwent complex surgery in 2021, specifically the structured and unstructured fields where information on health-related needs has been historically documented. We calculated how many hours case managers and social workers spent on resolving health-related needs, as recorded in a productivity flowsheet, and assessed how these needs were captured in the EHR.

### Phase I

#### Discover

Our research team conducted one-on-one interviews with care team members and patients from the Department of Surgery at our AMC. In these interviews, we aimed to identify current barriers and facilitators of collection and management of health-related needs information. Interviews examined three main areas of interest: (a) current workflows for pre-operative, peri-operative, and post-operative care; (b) current workflows and processes for collection of health-related needs information with a particular emphasis on social needs; and (c) participants’ preferences regarding the design of digital solutions to manage unmet needs. See Supplementary Files 1 and 2 for patient and care team interview guides, respectively. All study activities were approved by our AMC Institutional Review Board.

### Study Sample

#### Care Team Members

Care team members were recruited from our AMC Department of Surgery. Inclusion criteria included attending physicians, resident physicians, social workers, case managers, and advanced practice providers who work in the Department of Surgery and are at least 18 years old. A convenience sample was identified, and eligible care team members were recruited. All participants were consented to the study by a trained research team member before the interview. Each non-physician care team member received a $50 gift card after completion of both interview phases (i.e., discovery and design sessions).

#### Patients

Patients were recruited from our AMC Department of Surgery. Inclusion criteria were English speaking patients of at least 18 years old who underwent complex surgery (e.g. cardiovascular, bariatric, or gastrointestinal procedures) requiring at least one overnight stay. Patient recruitment was stratified by racioethnicity and neighborhood Ohio Opportunity Index (OOI) score (lesser opportunity and most opportunity), and eligible patients were recruited. We oversampled patients by racioethnicity based on evidence that indicates disadvantaged populations have less access to quality care compared to other individuals in surgery.^24,25^ One of these studies also noted that marginalized communities were more likely to experience adverse surgical outcomes.^25^ In addition to racioethnicity, neighborhood characteristics may also play a role in surgical outcomes and patient experience. An individual’s residence can affect their air quality, water quality, and transportation access which critically impact their health.^26^ Thus, we stratified patients by OOI, which is an area deprivation-based measure. The OOI is comprised of social and economic opportunities (i.e., income, employment, transportation, education) present across the state of Ohio. OOI scores range from 1-100. Higher OOI scores indicate greater opportunity in a neighborhood. Further details regarding the development and use of OOI can be found in the study by Fareed and colleagues.^27^ In our study, we divided OOI scores into septiles and used the septiles to define two groups: lower opportunity represented by neighborhoods with OOI scores that are in septiles 1, 2, 3 and 4; and higher opportunity represented by neighborhoods with OOI scores higher than the 4th septile.

### Semi-structured Interviews

One-on-one interviews were audio recorded and lasted approximately 60 minutes. The research team conducted the interviews using semi-structured guides. The interview guides were designed by the research team, pilot-tested on care team members and mock patients, and refined based on feedback from the pilot. The interview guide for care team members consisted of four sections: care team member background (roles and care workflow), medical care (typical course of treatment and challenges to medical and behavioral recommendations), collection and management of social needs information, and desired features in a digital solution for managing unmet needs. The interview guide for patients included four sections: patient background (surgery procedure), medical care, information on social needs, and desired features in digital solutions for managing unmet needs. Questions for the guide were based on prior research and guidance from experts in the research team.^28–30^. See Supplementary Files 1 and 2 for patient and care team interview guides, respectively.

Interviews were conducted virtually on Zoom or in-person at our AMC. All interviews were transcribed verbatim using the online platform Zoom. Primary interviewers were female in their early 20s and males in their late 20s and 30s. Secondary interviewers included a male in their early 20s. All interviewers were English speaking.

The research team discussed after each interview whether new themes emerged and achieved agreement for the themes of our study to ascertain thematic saturation. Codebook available from the author upon request.

### Define

We used a thematic analysis approach to organize care team member and patient interviews through open coding. Interviews were analyzed abductively: deductively to categorize results and inductively to discover emerging themes and subthemes. Initially, two team members coded three transcripts independently for each group to ensure our codebooks were appropriate. These team members staggered the coding process for the remaining transcripts. Team members met weekly to review the consistency of coding and agreement about code definitions. Team members also discussed the recurrence of themes to ensure data saturation was achieved. A third, senior researcher independently reviewed the coded transcripts and made final decisions when necessary. Two researchers selected exemplary quotes that reflected emerging themes from the transcripts. To maintain rigor, we performed peer debriefing and member checking with care team members to confirm our identified themes. We also documented our decisions during the data collection and analysis stages. Microsoft Word was used to manually code and analyze the transcripts. We followed COREQ guidelines in reporting our findings (See Supplementary File 3).^31^

### Phase II

#### Develop

For the second phase of our study, we asked participants about how to enhance patient activation levels in preparation for their surgery and recovery. Patient activation is an evidence-based concept that represents an individual’s confidence, self-management skills, and self-efficacy.^32^ Given our emphasis on whole-person health, we chose sample systems and tools that encompass health-related needs: medical needs (e.g., hypertension, diabetes), behavioral needs (e.g., diet, alcohol, and tobacco use), and social needs (e.g., food, transportation) to ensure a holistic and integrated approach to patient care during the surgery journey. In order to not assume that a patient is solely responsible for addressing their needs, we augmented our framework with social cognitive theory and goal setting theory,^33,34^ both of which assume the need for a supportive environment within which a patient can successfully feel activated and achieve their treatment goals. Our team utilized three Epic®-based products to gain initial feedback on available designs that can encompass a digital solution to our identified issues from Phase I. We chose to use Epic-based tools because of the familiarity of the system at our AMC among participants and to generate insights that can be generalized to other settings given the ubiquity of Epic among health care systems in the U.S. None of these tools were widely deployed at our AMC at the time of our study or were considered as part of the AMC goal of providing whole-person care for patients.

The three tools that were shown during the design sessions were Longitudinal Plan of Care™ (LPOC), Compass Rose™ (CR), and EpicCare Link.™ LPOC™ provides a detailed summary of a patient’s treatment plan, goals and active plans. This dashboard could be seen by care team members at our AMC to manage and guide patients on their goals. The main features on the LPOC™ dashboard are care coordination notes, allergy and problem lists, social determinants of health (SDoH) wheel, goals, and active plans. There are four main features of the LPOC™ patient view: goals, to do list, medications, and care team information. CR™ is a care coordination tool that allows the care team to make referrals for patients with social and behavior needs to community organizations (e.g., food pantry). CR™ offers features that allow the user to monitor steps necessary to achieve a specific goal (e.g., complete an application for a referral program). The main sections of CR™ are program specific targets, timeline, care team information, and a sidebar that describes the programs the patient is enrolled in, social needs the patient has, and different types of risk scores (i.e., admission to the emergency department, general risk score, and low patient engagement risk).^35^ EpicCare Link™ is a platform that allows community-based organizations (CBOs) to access care management tools. CBOs can access an Epic portal and message a care team member at the AMC, add to the patient’s LPOC,™ and help address patient care gaps. CBOs can update a care coordination note and update steps related to a specific patient’s goal that is subsequently integrated to the AMC EHR. The main features of the EpicCare Link™ dashboard are care coordination notes, allergy and problem lists, SDoH section, goals, care team, and recent visits.

#### Deliver

We conducted design interviews with a subset of participants from Phase I of our study. Care team members were shown an example action plan for a patient who needed to make sure they had a high protein diet for six weeks post-surgery. This action plan was conceptually organized with a specific, measurable, achievable, relevant, and time-bound (SMART) goal. SMART goals can guide patients to achieve a clear and tailored health goal.^36^ Care team members were then shown three screenshots of digital tools: LPOC™, CR™, and EpicCare Link™ depending on their role at the AMC. The clinical care team were primarily shown LPOC™ and CR.™ The non-clinical care team were primarily shown CR™ and EpicCare Link.™ Patients were shown the same example of the action plan as the care team and the patient view of LPOC.™ Care team and patient participants were also asked about whether the example prototypes will improve patient activation levels. See Supplementary File 4 for sample SMART goal and patient action plan.

To analyze the design interviews, we conducted a sentiment analysis based on features. Codes were created based on specific features of the digital tools that were presented during the design sessions. A researcher manually coded all Phase II transcripts using MAXQDA® and subsequently conducted the sentiment analysis in MAXQDA to categorize quotes as positive, slightly positive, neutral, slightly negative, or negative sentiment. This approach allowed our team to determine the nuances of user perceptions and attitudes on the digital tools and identify specific features that enhanced whole-person care for complex surgery patients. The MAXQDA sentiment analysis feature uses the SentiWordNet 3.0 lexicon for English.^37^ We selected the stop word list option on MAXQDA to ignore words that do not have any sentiment. After this step, two members of the research team manually reviewed the MAXQDA codes and overrode miscoded quotes. Another researcher reviewed the manual and MAXQDA codes to ensure that the coding was accurate and consistent. A senior researcher selected exemplary quotes for each sentiment. We evaluated the digital tools based on sentiment, and cataloged the desired features and recommendations provided by participants for a final set of desired user specifications in a whole-person based digital solution.

## Results

### Participant characteristics

We recruited 20 care team members and 24 patients for our study. Details about care team member and patient characteristics are noted in Tables 1 and 2, respectively. Care team members consisted of 35% physicians, 20% resident physicians, 20% case managers, 15% advanced practice providers, and 10% licensed social workers. Eight care team members were male, and twelve care team members were female. The average age of care team members interviewed was 41 years. Among patients who were interviewed, 42% were NH white, 46% were NH Black, 8% Hispanic and 4% NH other. Among the sixteen patients living in a lower opportunity neighborhood, 38% identified as NH white, 44% identified as NH Black, 13% identified as Hispanic, and 6% identified as NH Other. Among the eight patients living in a neighborhood with a higher opportunity, 50% identified as NH Black, and 50% identified as NH white. Thirteen patients were male, and eleven patients were female. The average age of patients interviewed was 59 years. Approximately 63% underwent gastrointestinal/benign surgery, 8% underwent gastrointestinal/oncology surgery, 13% underwent general/hernia surgery, 4% underwent general/soft tissue surgery, 8% underwent cardiovascular surgery, and 4% underwent non-gastrointestinal/oncology surgery. In Phase II, we conducted digital sessions with a subset of participants from Phase I. We interviewed a total of nine care team members: 33% case managers, 22% APPs, 22% social workers, and 22% physicians. The nine care team members were female. We interviewed a total of five patients, of which three patients were female and two were male. The average age of the patient participants was 58 years old. Sixty percent of patient participants underwent gastrointestinal surgery and 40% underwent general surgery. Among patients interviewed, 80% were NH white and 20% were NH Black. Among the patients who identified as NH white, 50% live in a neighborhood with a lower opportunity. The patient who identified as NH Black lives in a neighborhood with a lower opportunity.

**Table 1.** Care team member background information.

| Characteristic | Distribution (N = 20) |
| --- | --- |
| **Role (n, %)** |  |
| Attending Physician | 7 (35%) |
| Resident Physician | 4 (20%) |
| Case Manager | 4 (20%) |
| Advanced Practice Provider | 3 (15%) |
| Social worker | 2 (10%) |
| **Gender (n, %)** |  |
| Female | 12 (60%) |
| Male | 8 (40%) |
| **Age (mean, SD)** | 41 (±12) |

**Table 2.** Patient background information.

| Characteristic | Distribution (N = 24) |
| --- | --- |
| **Racioethnicity (n, %)** |  |
| NH white | 10 (42%) |
| NH Black | 11 (46%) |
| Hispanic | 2 (8%) |
| NH Other | 1 (4%) |
| **Ohio Opportunity Index (OOI)** |  |
| Lesser Opportunity | 16 (67%) |
| Most Opportunity | 8 (33%) |
| **Gender (n, %)** |  |
| Male | 13 (54%) |
| Female | 11 (46%) |
| **Age (mean, SD)** | 58 (±17) |
| **Surgery type (n, %)** |  |
| Gastrointestinal/benign | 15 (63%) |
| Gastrointestinal/oncology | 2 (8%) |
| General/hernia | 3 (13%) |
| General/soft tissue | 1 (4%) |
| Cardiovascular | 2 (8%) |
| Non-gastrointestinal/oncology | 1(4%) |

NOTE: OOI categories were defined as follows: lesser opportunity OOI which includes septiles 1 through 4, and most opportunity OOI which includes septiles 5 through 7. Abbreviations: NH = Non-Hispanic.

### Diagnosis Phase

#### Problem

Surgery patients and their care team members struggled to effectively collect and manage health-related needs, particularly social needs. Care team members faced challenges such as time constraints and the inability to identify appropriate resources for their patients, which can delay patients’ ability to achieve clinical treatment goals. Patients, moreover, are overwhelmed with social needs while trying to balance these needs with preparation and recovery from their surgery.

Based on the current literature, care team members faced challenges with addressing health-related needs, particularly social needs due to time constraints.^20^ Care team members felt that they do not have the time and resources to successfully address these needs.^38^ We found that case managers and social workers who recorded their activity time in the EHR productivity flowsheet spent an average of approximately six hours per patient addressing social needs. Some care team members offered a resource packet to patients. Social Worker 2 explained:

> *“Yeah, so we feel like, it’s kind of a false narrative. Some people think that we can like get them an apartment which, unfortunately we can’t. I don’t really have the time during the day. Unfortunately, so, if they’re homeless, we have like a homeless hotline. We give them and then for the housing resources we have lists of like section 8, housing and apartments. That we give to the patient and kind of just say I’m not sure like what’s available for these departments. But here’s like kind of a starting point but it’s kind of it, or like a lot of people have like eviction issues which we try to do as much. We can’t really do much with that. Unfortunately, I just give them the resource packet and say, here’s some organizations that might be able to help you. I try not make any promises, because some of these organizations unfortunately run out of money pretty quickly.”*

Care team members shared that common challenges for patients to meet medical goals are lack of access to resources. Case Manager 1 explained:

> *“I would say the big ones tend to be like lack of support, whether that’s if they don’t have a lot of support people, or they don’t have transportation or, you know, lack of resources. If you know their lower income or are not insured or under insured. Those are obviously big barriers and then additionally, more kind of generally on the medical side, would just be general compliance. which sometimes can just be an education gap, and other times is just patient preference will say, like, if they’re just kind of not doing or following the intended care plan.”*

There are several challenges that prevent patients from meeting their medical goals. These challenges differ depending on the population.^39^ Physician 1 mentioned:

> *“I think there’s, I mean, clearly, there’s social determinants of health and kind of health literacy components social support, you know, and family members or loved ones who are able to encourage adherence to plans and treatment strategies. Certainly, in the bariatric population there’s a lot of metabolic component to their adherence to you know, durable weight management and diet patterns. Obviously, there’s financial problems. But that’s kind of encompassing what we talked about. But you know, I think travel can be challenging for our patient population, who sometimes is coming in from out of state just to see some of us. Yeah, those are the ones that come to mind.”*

For example, patients may experience financial issues which can be a major challenge in following care team recommendations and may delay medical treatment.^40^ Patient 23 shared:

> *“Yeah, she just told me I needed plastic surgery. Stuff like that is, her recommendation is to do the plan to switch it, because I’ve been coming to the hospital for like maybe three times they do surgery about the same armpit, and she says, you recommend me to do plastic surgery. And I say, Okay, I will talk to them, man. It’s hard for me. You know what I mean. I’m working for Uber. You know. I’m driving. I don’t know whether I will qualify to collect disability. You know what I mean? Because that’s it’s not a small surgery. It’s a big surgery that would take like maybe three months to stay home, and I have family to feed. I got kids. I got mortgage to pay. So it’s crazy. I have to plan that. It’s not something I can just come do it. you know, doesn’t work like that. And he referred me to the process. So you to talk with them. And I said, Okay, I will do that. And now I have a referral to go there to talk to them. So that’s what we’re saying. She’s the one who will do the surgery, so she look at it and see that. Oh, yeah, look good. You know what I mean. He put the band aid back again. I said, this is, look good. It’s coming, you know. Start getting close. Sounds good.”*

### Digital solution tools

Digital health advancements provide an opportunity to better address health-related needs in the hospital system. A closed-loop approach has been proposed in the literature for addressing these needs, with communication, management, and resolution between the patient and care team. As previously noted, we used three Epic® tools that could be fully implemented at our AMC to potentially align with this approach. We coded user feedback during the design interviews and organized quotes based on sentiment. For the care team, EpicCare Link™ had the highest number of quotes (25) with positive or slightly positive sentiments. LPOC™ and CR™ each had 17 quotes coded as positive or slightly positive sentiment. The patient view of LPOC™ had eight quotes with positive or slightly positive sentiment. Care team members saw the value in incorporating these digital tools into their current workflow. Overall, the care team understood the importance of most sections in each digital tool. However, there were suggestions to add customizable features which yielded a slightly negative or negative sentiment such as adding more information or adding a drop-down option for a section. CR™ had the most quotes (19) with slightly negative or negative sentiment. EpicCare Link™ had the second highest number of 17 quotes with slightly negative or negative sentiment. LPOC™ had a total of 14 quotes with slightly negative or negative sentiment. The patient view of LPOC™ had five quotes with slightly negative or negative sentiment.

Patients overall liked the idea of having a digital tool designed for their health-related goals on the patient view of LPOC.™ There were recommendations made that focused on re-wording sections to encourage patients to be more engaged in their active plan. There were 21 quotes coded with slightly positive or positive sentiment. There were 23 quotes coded with slightly negative or negative sentiment for the patient view of LPOC.™

### Phase I

#### Discover

When we asked care team members about health-related needs, many expressed the need for local resources that address social needs and for a streamlined process to collect and manage this information. When we asked patients about health-related needs, they expressed social needs challenges and emphasized the importance of closing the loop to resolve these issues. Our interviews captured barriers, facilitators, experiences, and future considerations with health-related needs

##### Perceived Impacts of Managing Health-Related Needs

###### Benefits

Both care team members and patients understood the value of the collection and management of health-related needs information. Care team members emphasized that health-related needs can affect the patient’s surgical outcomes and their overall health. Case Manager 4 mentioned:

> *“Honestly, it can make or break the success of the overall care because you can say: Hey, we’re gonna do this as a doctor. We’re gonna do this, this and this cause that. What that’s what needs to be done. If they don’t have food to heal that wound. It’s not gonna heal right if they don’t have clean water. If they don’t have transportation or insurance information or somebody that can help them. Then they might have a setback like I said, in the perfect world everything I listed out for you on that piece of paper or that care plan is smooth. That never happens, you know that. But there’s a ton of factors that we try to address even before they leave the building, so that if there is something that I’ve anticipated potentially could come up. This is the resource that you need to call for that make sense.”*

Patients felt that the collection of health-related needs information can help them better prepare for their surgery and recovery process. Patients believe that feedback on health-related needs from the care team is important and feels like they are being cared for. One patient shared that the collection and management of health-related needs information is necessary to effectively capture and assess patients’ health. Patient 7 shared:

> *“So inside, it’s a necessity. You’re not trying your best. If you’re not doing that. Okay, you know you’re shorting yourself. All you’re doing is you’re shorting yourself. You don’t give them the full story.”*

###### Barriers

Care team members believe some patients may not be comfortable or honest with sharing health-related needs information. Patients may share information, but do not want the information to be on their record (e.g., domestic violence). Social Worker 2 shared:

> *“If patients are forthcoming. Sometimes they’re embarrassed. They don’t want to talk about it, which I totally understand. But it just depends. Like, I said, some patients there are patients that need a lot of things, but they just don’t want to talk to me. I think sometimes the social worker can leave a bad taste into that. Some people’s mouths. They might have had interactions with social workers in the past, and it been kind of negative. So it just depends on who the patient feels comfortable talking to. I feel like that’s a big one as well.”*

A patient shared during their interview that they would not feel comfortable sharing health-related needs information, particularly social needs, with their care team. Patient 19 explained:

> *“[TRANSLATOR SPEAKING] So, he was basically saying that like he didn’t get like, why, it would be important to like share your social needs with the doctor, because he says like, even though he would have problems, he wouldn’t specifically communicate it with doctor…He just wouldn’t feel comfortable telling. It’s out there like he needs this, or he needs that. He just doesn’t feel good… He just feels like sharing those needs isn’t something that he wants to do or he would like to do. He doesn’t feel comfortable.”*

Another patient felt like a burden to the care team with their health-related needs and as a result did not want the care team’s help with addressing them.

One patient highlighted that the health-related needs information is being collected but not managed properly. There is a lack of feedback to patients to address these needs. Patient 20 shared:

> *“I think a lot of the information goes in, and that’s [IT] and it just stops.”*

It was mentioned that patients with lower socioeconomic status may have difficulties that the care team must consider during the collection and management of health-related needs processes. Another patient emphasized that the collection of health-related needs information was redundant and questioned why this information is not synthesized in one place for the care team to reference. Patient 7 noted:

> *“You know, I’ve been asked a lot of the same questions over and over again. So I was wondering why it wasn’t all documented in my charts, so everybody could see the whole story. My story, and as thorough as I find my charts to be. and I really like that program. I found it a little bit redundant, but at the same time I was very patient about answering all the questions when it was asking.”*

##### Current Collection and Management of Health-Related Needs

###### Awareness

Care team members were aware of the collection of health-related needs information, it was not consistently implemented. Resident Physician 1 stated:

> *“Yes and no [PATIENTS REPORT HEALTH-RELATED NEEDS]. I mean, I think if we ask if we ask about it, and we ask about it because they’re usually relevant to their condition. Yeah, I think they certainly do report it. You know, unless we’re asking about it. I don’t think it’s like freely brought up often.”*

Those who do not collect information may informally ask their patients questions relevant to their practice (i.e., asks a patient whether they have social support for recovery post-op). Physician 5 shared:

> *“So they don’t frequently offer it. No, it’s not something that is routinely part of our assessment, really. And I always ask people the things that I do always ask, are you gonna be able? Are you gonna have somebody that’s gonna be able to help you out afterwards? But I don’t go into the nitty, nitty gritty of the social situation. I’ve had that bite me a couple of times.”*

The Epic SDoH Wheel^®^ is available to care team members at our AMC. We found that many care team members were not familiar with this feature. However, care team members may use other Epic^®^ functions for noting health-related needs (i.e., Inbasket messages or progress notes in Epic^®^). Advanced Practice Provider 1 shared:

> *“I’m not sure what that is the social determinant of health wheel. I’m not sure. I mean, I use the in basket all the time. And you know, for calls and messaging like my chart messages. We use that pretty much every single day. But I’m not sure what the social wheel is.”*

Many patients could not recall if health-related needs information was collected before, during, or after their surgery. Some patients recalled being asked once a description of social information was provided in the interview. Other patients were unfamiliar with health-related needs. Patient 6 explained:

> *“Like I said. You know I was there a couple of 3 days. I said I need a shower, and they’re very obliged, and they got me hooked up. We wouldn’t, you know, have shower and everything. So I mean That’s social needs, cause I was feeling pretty funny, and I, you know. But they hooked me up.”*

###### Roles

Some patients could not recall names of care team members who collected their health-related needs. Patients reported that case managers were the care team members most involved in the collection of health-related needs. This aligns with what care team members shared. Care team members that collect health-related needs ask for both medical and social-related needs. Case managers and/or social workers are the specified team members responsible for the collection of this information. Social Worker 2 noted:

> *“So for all patients that like have any needs like homelessness, addiction, medicine like issues of transportation, etc. We’ll get a consult from the nurse to do like our social work assessment, or I’ll just do a chart review and kind of see that their needs and so I’ll complete our social work assessment, just asking some questions about the social determinants of health and then giving. We have, like a really big resource packet. We give any specific resources for like housing and things like that.”*

###### Documentation Process

A psychosocial assessment – a structured tool – is used by case managers at our AMC to collect some health-related needs information. This information is placed in the EHR progress notes for all care team members to review; however, only few review it outside of case management and social work. Care team members who provide oncology care use the Oncology Distress Survey to collect health-related needs information. Advanced Practice Provider 2 shared:

> *“There is the [ONCOLOGY] distress screen. Are you familiar with that? So that gets sent to all of the oncology patients? And then if they fill it out on my chart and it flags, it comes back and the flow is that you’re supposed to call the patient. If there are flag responses and discuss referrals.”*

Other care team members evaluate where the patient currently is before surgery regarding health-related needs (e.g. employment, financial issues) to assess peri-operative risk and use this information to develop a surgical plan. They have their own questionnaire and ask questions verbally. The collection process varied greatly among patients. One patient mentioned that they had small talk with a care team member. Other patients recalled a questionnaire that focused on food and housing. Patient 21 noted:

> *“It was a series of questions that they would ask on the screen. There were three different people that come in and ask the same question one after the other.”*

Some care team members expressed that they are uncomfortable to ask patients direct questions regarding health-related needs. Physician 1 mentioned:

> *“I ask everyone some social questions. You know. To me what’s most important is their smoking status and smoking history. And I asked everyone about their work or their overall day to day activity levels. I think that’s important in a surgical practice. I sort of intentionally do not ask them a lot of other social things that could be that can be a little off putting to patients, I think, who are simply requesting a surgical procedure be performed. And a lot of times, I think. If they want to volunteer information. Talk about information I’m happy to. But, for example, I don’t take a detailed sexual history or a detailed religious history or you know, maybe even just a relationship. History is not of interest to me, because I don’t think I’m going to impact that for the patient dramatically. And if they have questions about that, that I’m happy to talk about it. But so I was asking about their work. And I say, what kind of work do you do? What kind of work did you do? And I feel that that’s very important for the patient relationship? you know, just in terms of me getting to know them and their family, and how I can help them best. And you know II regularly type in my note. This is a [in their 40s] avid golfer, you know, because to me, if I’m going to be fixing their abdominal core musculature, I need to think about the fact that this guy golfs every day. That’s gonna impact the way I do surgery or what technique I use or what long-term outcome. I’m hoping for that patient which is different than somebody who’s wheelchair bound, for example. And maybe it’s, you know, retired whatever. So I always ask those questions.”*

This collection process may be seen as redundant. Another patient mentioned that the care team member documented health-related needs but did not follow up or use that information. Patient 20 shared:

> *“No, they came to solve me, and I gave them the same information. But and they supposedly well, but not, there’s no follow up…They just asked it and annotated it. And that was it.”*

###### Monitoring health-related needs

Some care team members have a unique way of monitoring health-related needs. For instance, in the note history, a care team member will place a plus sign to indicate that they had a conversation regarding a sensitive topic the patient brought up. They can then refer to the conversation based on what the care team member recalls.

Care team members have noticed an increase in documentation of health-related needs. As a result, these care team members make the effort to review patient health-related needs. Case Manager 3 explained:

> *“Within the chart, the EHR chart. We’ll go and review, like what information’s available from progress notes and things that the social workers and outpatient center, or whoever else. Because I’ve noticed lately that more providers are being involved in that and asking those questions. So I will review that I always try to review those kind of things when the patients are coming in, so that I know kind of what we’re facing and what’s been done.”*

Some patients expressed that if they had a health-related needs issue, they would feel comfortable enough to let their care team know. Patient 5 noted:

*“Yeah, I would at least call up either by calling them in or sending a message through like a portal.”*

###### Basic social needs

We defined a basic social need as a need that affects the patient’s daily life, but does not require a comprehensive solution from a financial perspective (e.g. food, transportation, medication assistance). ^41,42^ Transportation was a basic social need that was mentioned most frequently. Food insecurity and medication assistance were the second and third most frequently mentioned basic needs among patients, respectively. Transportation may be the highest because many of the AMC patients travel commute from long distances to the AMC for care. Social Worker 2 shared:

> *“Yeah, I would say transportation to honestly a big one that I didn’t, that we run into a lot. But the patients have rights home can’t drive when they leave here. So I think transportation is a big one, and just making sure that patients have a safe ride home.”*

Some patients live far from the AMC which can contribute to transportation issues. One patient wished they had known about reporting transportation issues to the care team. Patient 14 shared:

> *“That would be excellent [RECEIVE TRANSPORTATION ASSISTANCE], because if I could have reported transportation problems, my son could have been at work today, and their bus could pick me up, brought me here took me up. That would have been perfect.”*

###### Complex social needs

We defined a complex social need as a need that requires extensive assistance with different factors of patients’ daily lives. This involves a more comprehensive solution from a financial perspective (e.g., housing, finances, employment, education, social support and home care, air quality, domestic violence).^41,42^ Based on our interviews, social support and home care was the complex need with the highest frequency. Social support and home care include patients who may live alone or lack friends or family to help with their care and recovery.

Among patients, the most frequently mentioned complex need was social support and home care. Some patients needed someone at home to change their dressing post-surgery. One patient shared that they lived alone and had a family member stay with her to make sure someone was near her during recovery. Patient 2 shared:

> *“So I do have issues, and I and I do live alone. I didn’t mean to. I don’t know if you thought someone live with me, but I really don’t have anyone that lives with me…Yeah [I had someone stay with me after surgery], just actually, the first night that I came home. Yeah, my cous, my cousin was here, but she had a dog, which the dog irritated me, so I said, I don’t think I need you, I said I. My arms aren’t broken and my legs worked fine, so I don’t think I need anyone. I feel fine. I felt fine when I came home is really very strange for the surgery that I had. So I didn’t expect to feel so well afterwards. But yeah, I did.”*

Home environment/home safety and financial concerns were the second and third highest complex needs, respectively. Figure 1 provides a breakdown of basic and complex social needs for care team members and patients. Among patient participants who discussed social needs when we specifically inquired about basic and complex needs, 75% (n=9) lived in lower opportunity neighborhoods, approximately 50% of whom identified as NH Black. We also found that >50% of patient participants who did not discuss any social needs during our interview lived in lower opportunity neighborhoods, over 50% of whom identified with a marginalized community.

**Figure 1.**
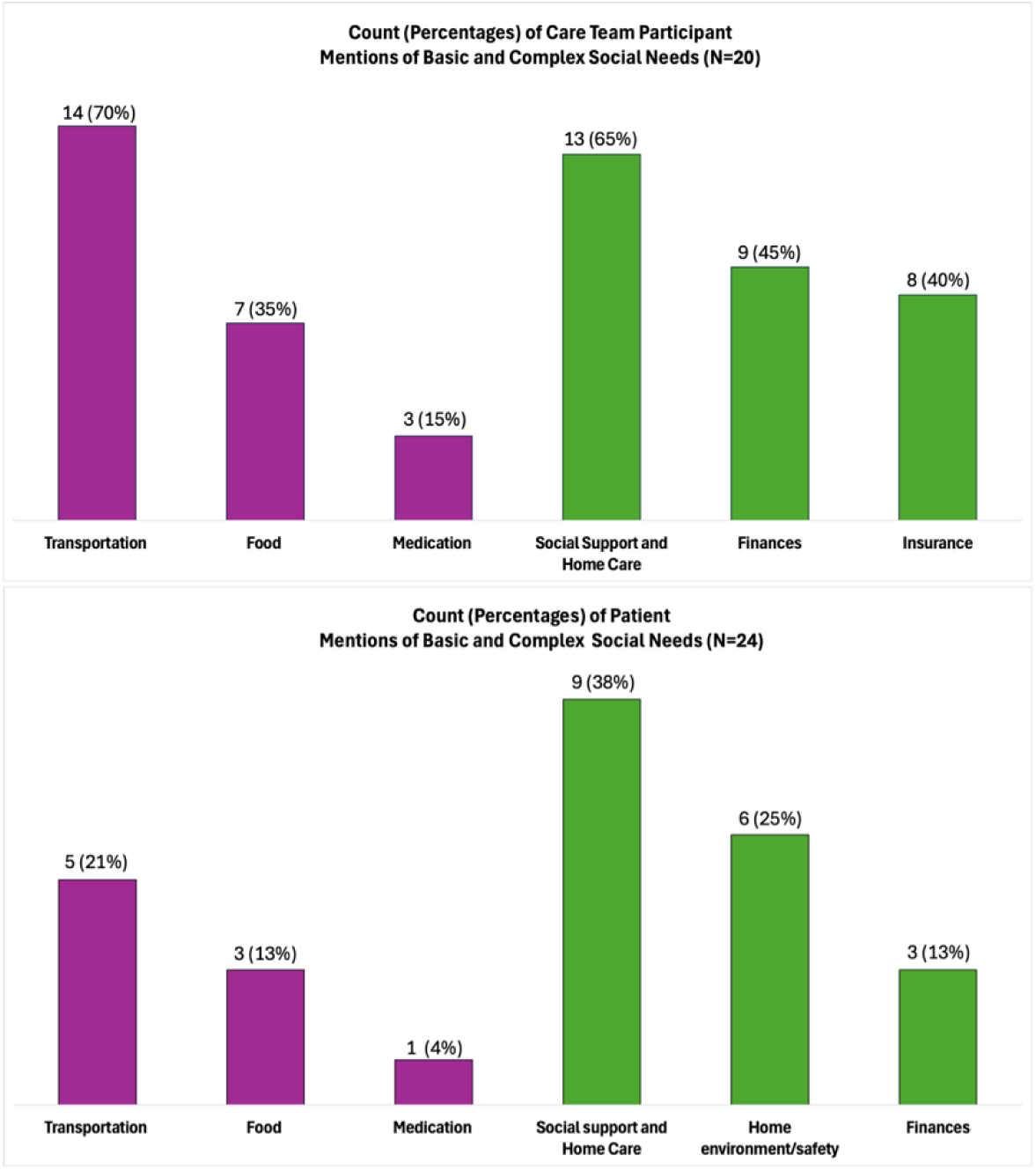
Frequency of the top three basic and complex social needs for both care team members and patients Percentage of mentions of basic and complex social needs among Panel A: Care Team Participants and Panel B: Patients. Basic needs (purple) include transportation, food, and medication; complex needs (green) include social support/home care, finances, insurance, and home environment/safety

###### Resolution of health-related needs

One care team member specifically shared that they did not know where to refer patients to address a social need. Care team members focused on medical needs may have the patient connect with a care team member centered on social needs (i.e. social services, social worker, case manager). Some care team members noted that they are not familiar with local resources in their patient’s communities, especially rural areas. Physician 2 noted:

> *“Usually what I do is I loop in social work or the PCRM [PATIENT CARE RESOURCE MANAGER] to help with that? Because although a lot of my patients are here over in the [DEIDENTIFIED] community. A number of them come from farther away, and I don’t know the resources for the cities that are like two and three hours away. So sometimes I’ll identify it, and then try to get them looked in with somebody who can be a little bit more boots on the ground to turn in terms of understanding that because that’s kind of outside of like what I know of the community.”*

Recently, the AMC implemented the UNITE US^®^, a platform integrated with Epic^®^ intended to provide automated access to resources. Some care team members expressed concerns about “closing the loop” with patients to ensure that they have indeed received the resources they were referred to due to challenges such as inadequate community capacity and linkages for referrals made through UNITE US^®^. Physician 5 explained:

> *“We need to support? The answer can’t just be that the patients are. Gonna tell us more about their problems. The answer needs to also be that we have solutions to those problems. So my experience when patients do bring up those things is, I’m very much at a loss. I say, wow! That’s horrible. I’m really sorry that you’re going through that. I can’t. When a patient comes to me and tells me that their husband is unwilling to help them with their recovery, because he’s out drinking every night. What? What do I? What I do with that actually had that happen recently? And I asked her and made sure she was safe. There was no concern for abuse or anything like that. But yes, she felt really, really lost, and really unsupported, and what I can’t, what can I do about that? So asking people to report is one step, but being able to support is a separate and I think more important. Step right? So if you ask people things and then don’t provide solutions. They feel like you’ve just given like, why did you even ask. People don’t react to that. Very well.”*

Patients experienced some challenges with resolving health-related needs. Some care team members did not follow up with a patient on a health insurance related concern. Patient 20 shared:

> *“You know I put this question out several times. It’s not that we need it. But I wanted to get my wife reimbursed, so let me explain. There’s a place, I guess, an agency, and I haven’t looked at. It’s called [DEIDENTIFIED STATE] job family services, or something like that. And this agency supposedly reimburses individuals. Family members for the help. But my wife is doing a fantastic job, and I pay on myself. But it’s the same money. So I said, let’s see if you can get. So a friend of mine said, Hey, Christine can get reimbursed for her time and effort. I’m like, really, okay. So they said, call them. I’ve asked that question four or five times through my care team. I’ve never got a response to that question of who should I go through? Is there a telephone number, and I know this is not the 1st time that the career team and the hospital has heard that you know how what agency reimburses for that I know some. I guess it’s called Medicaid. And I know I don’t qualify for that, you know, and rightfully so, I mean, you know. Cause they said, It’s $24,000. I thought it was a month that you make. And I like, Oh, yeah. And then they said, No, sir, it is for a year, I’m like, Oh, no, I make a little more than that. And so you know. But there needs to be something like that in place where somebody follows up. If you, if a person has a question. Don’t just take down information, follow up either in my chart, which is a very expedient way, my chart, or send a person a message, or you have their numbers. Give them a call.”*

Another patient needed transportation for an appointment and the care team provided transportation assistance to the patient. A patient did not expect health-related needs to be addressed during the collection process. The patient thought the care team wanted a baseline of health-related needs history for patients.

See Supplementary File 5 for exemplary quotes from care team members and patients.

##### Future Approaches to Collect and Manage Health-related Needs

###### Define

We further analyzed the formative interviews using swim lane diagrams. A swim lane diagram allows us to visualize the interactions and workflows in the AMC. We used the “as-is” swim lane diagram to map out the current workflows for a surgical patient (Figure 2). We included workflows for the surgical patient, referring physician, advanced practice providers, surgeons, case managers, and social workers. Each role is indicated in a different color. The arrows and lines from one note to another represent the flow of interaction. Each note is color-coded to describe barriers identified by participants, possible digital solution features, questions, decisions to be made, and follow-up questions. We identified four critical gaps in our “as-is” swim lane diagram: (1) heterogeneity in the approach to screening, monitoring, and managing health-related needs; (2) patients may feel uncomfortable reporting health-related needs, particularly behavioral and social needs, to their care team; (3) lack of access to referral resources to resolve needs; and (4) the need for a closed loop intervention for patients and care team members. As previously noted, the care team spent almost six hours per patient on coordination of health-related social needs based on our assessment of patient EHR charts from 2021. We also identified some health-related needs collection in the EHR among 55% of patients. Most of the recorded needs were focused on tobacco and alcohol use, as well as depression. There was some data recorded in a Psychosocial Assessment structured field under clinical progress notes and some information was documented under social history.

**Figure 2.**
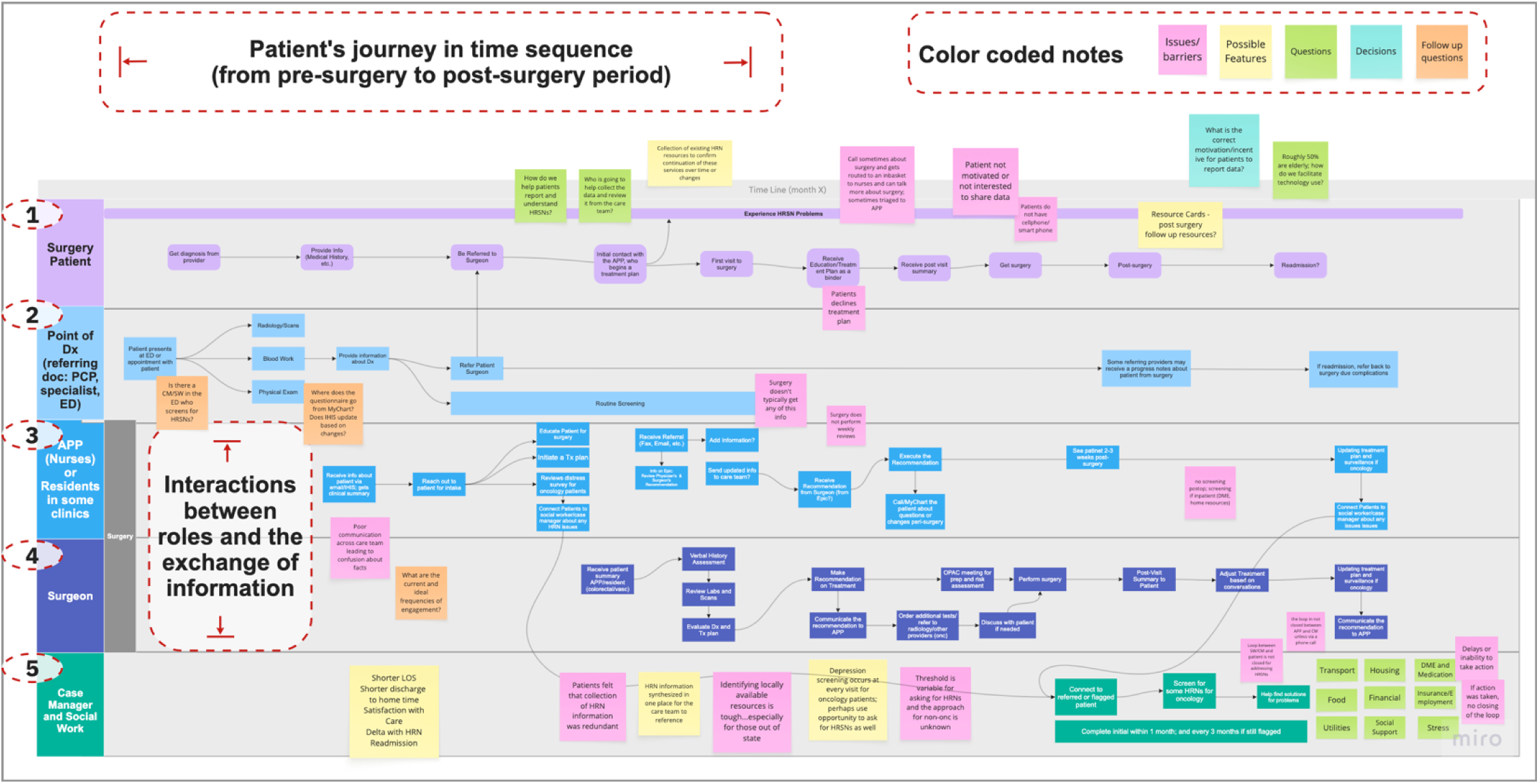
Swim lane of “as-is” clinical workflow Figure illustrates time-sequenced interactions across five roles: 1) Surgery Patient, 2) Point of Diagnosis, 3) Advanced Practice Providers (APPs) or Residents, 4) Surgeon, and 5) Case Manager/Social Work. Figured scaled to illustrate complexity of the system.

To address these critical gaps, we centered our design proposition on patient activation because the challenges a patient faces with regard to their confidence, self-management skills, and self-efficacy are associated with a successful complex surgery outcome. We further augment the need to enhance a patient’s activation levels with support from their care team that can be informed by other behavioral theories: Social Cognitive Theory (SCT) and Goal-Setting Theory (GST).^33,34^ SCT posits that successful performance of a behavior is dependent on an individual’s capabilities, cognitive influences, and environmental factors. These influences are organized in three domains: 1) knowledge and beliefs; 2) skills; and 3) self-efficacy.^33^ GST proposes that motivating individuals to enhance performance and improve self-efficacy requires goal clarity, commitment, feedback, and task complexity.^43,44^ Patients may not see the relevance or trust the care team regarding health-related needs and could be unaware of how addressing unmet needs can help them achieve their treatment goals. Based on GST, for example the surgery treatment plan can be developed as a playbook to ensure that the patient achieves their treatment goals. For patients to be motivated to achieve their goals, SCT is essential to provide the necessary information and skills for patients to take an active role in managing their health while being cognizant of their health-related needs, which can be acquired with support from the care team and the community.

### Phase II

#### Develop

Based on our results from Phase I, the research team created a “to-be” swim lane diagram (Figure 3) that addresses the identified major gaps of the current workflow. Given that health-related needs screening can involve sensitive topics such as housing or financial troubles,^29^ we implemented measures in our intervention to mitigate potential discomfort and help foster trust. Patients can report health-related needs using a web-application – accessible through a smartphone or the web, both independently or with the help of a nurse navigator. This standardized approach enables collection of health-related needs from a broad panel of patients. Once patients complete their health-related needs assessment, they will be automatically referred to resources to address their needs – triaged by the nurse navigator to specific care team members based on the type of need. Care team members will then follow-up on patients to ensure health-related needs have been met, with the intent to close the loop between the patient and the resource need.

**Figure 3.**
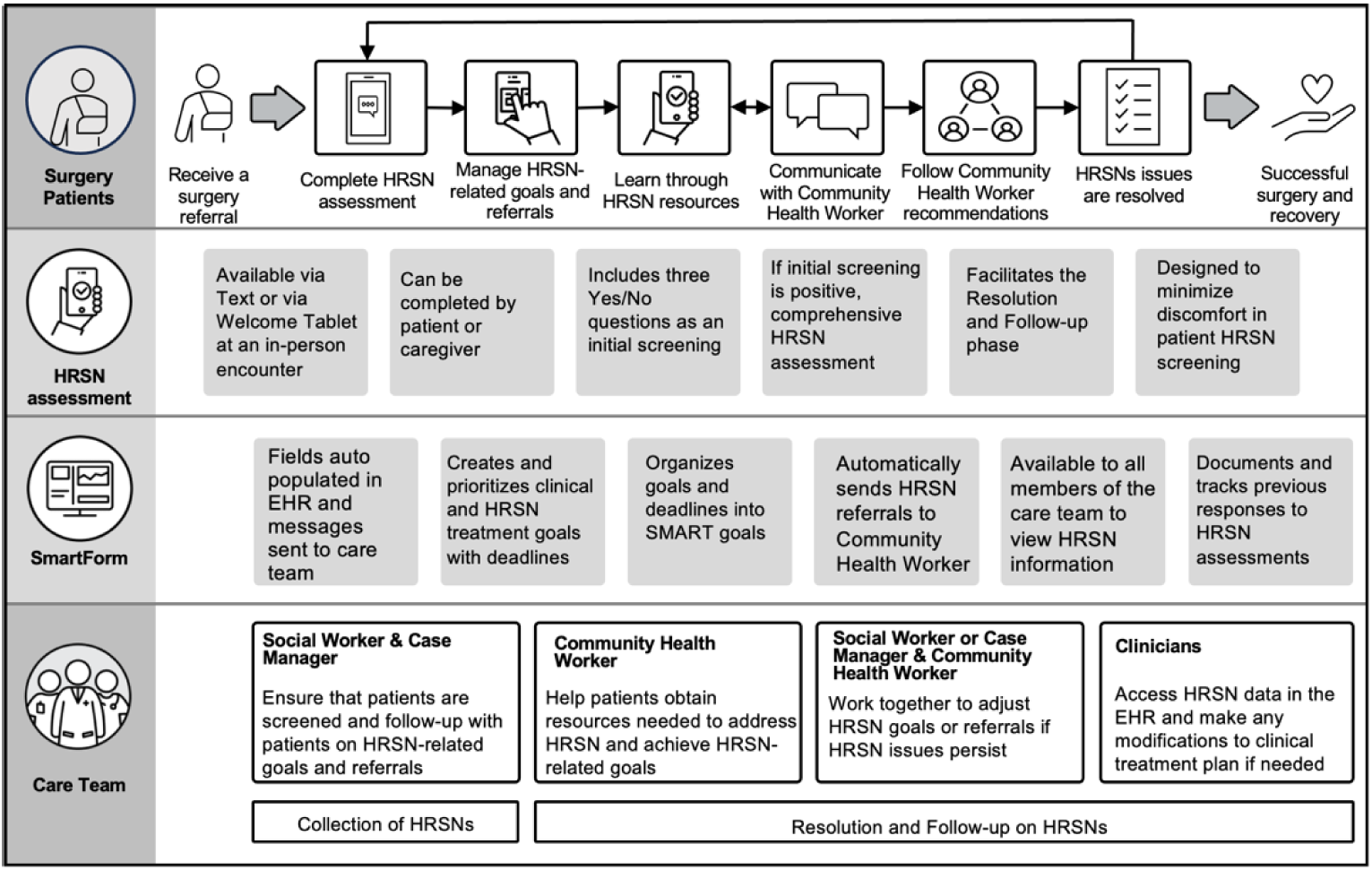
“To-be” swim lane of clinical workflow Figure outlines the process for identifying and addressing health-related needs among surgery patients

#### Deliver

In addition to an ideal workflow, we also gathered desired features for a digital solution that would be embedded in this workflow. Participants were asked questions during the design interviews about whether Epic^®^ tools would improve patient activation levels. Most digital tools had positive sentiment among the care team participants. LPOC,™ LPOC™ patient view, and CR™ had more positive or slightly positive sentiment than negative or slightly negative sentiment. There were equal number of quotes coded as positive or slightly positive and negative or slightly negative for EpicCare Link.™

Sentiments varied depending on the digital tool shown and tool specific features. The care team had the most positive or slightly positive sentiment (29%) for the active plans feature of the LPOC.™ The SDoH wheel had the most negative or slightly negative sentiment (43%). Three care team members expressed that they would prefer a list or tab section. The ability to print the LPOC™ was the feature with the most positive or slightly positive sentiment (50%) in the LPOC™ patient view. The to do list in the LPOC™ patient view had the highest number of coded quotes with negative or slightly negative sentiment (60%). The care team wanted to ensure that to do list items were tailored to the upcoming surgery and recovery. Referrals, social drivers, and the sidebar in the CR™ dashboard had the most positive or slightly positive sentiment (24%). The CR™ targets had the most negative or slightly negative sentiment (42%). Care team members wanted more information than what was shown to better understand patient’s needs specific to programmatic goals. The SDoH feature in the EpicCare Link™ view had the most quotes with positive or slightly positive sentiment (24%). However, the SDoH and goals features had the most quotes with negative or slightly negative sentiment (24%). Care team members wanted to ensure that the SDoH covered all social needs domains (e.g., financial and employment). The goals section needed to be clearer on the specific goal and progress towards that goal.

Patients overall felt that the LPOC™ patient view would improve patient activation levels and be helpful in preparing and recovering from the surgery. Based on the patient participants, the care team, goals and to do list features had the most coded quotes with slightly positive or positive sentiment (35%) for the LPOC™ patient view. The goals feature had the most quotes with slightly negative or negative sentiment (40%). Examples of recommendations from patients were including a progress on goals feature and providing alternative approaches to achieve goals. Tables 3 and 4 provide user specifications by feature and technology from the care team and patient design interviews, respectively.

**Table 3.**
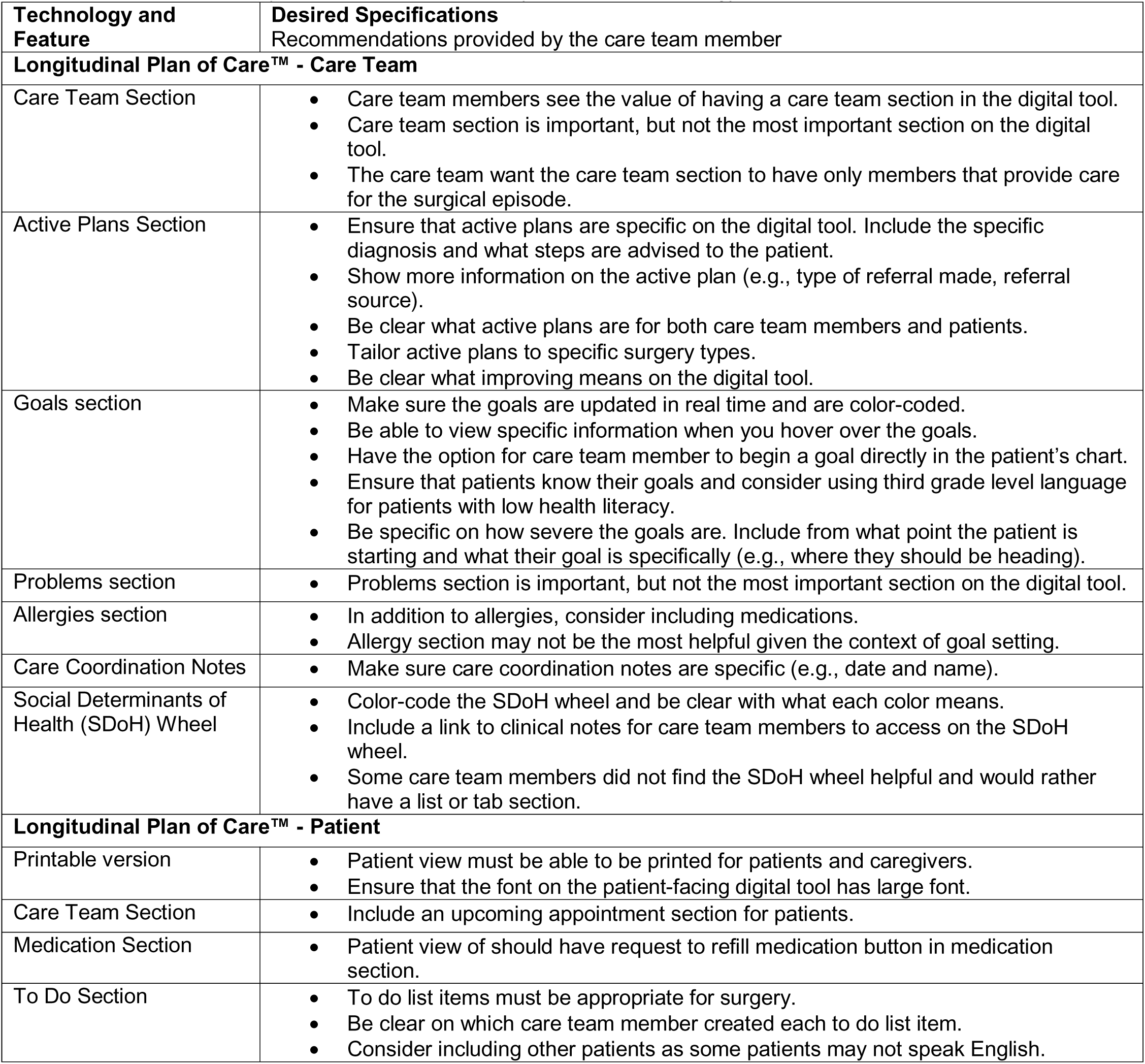
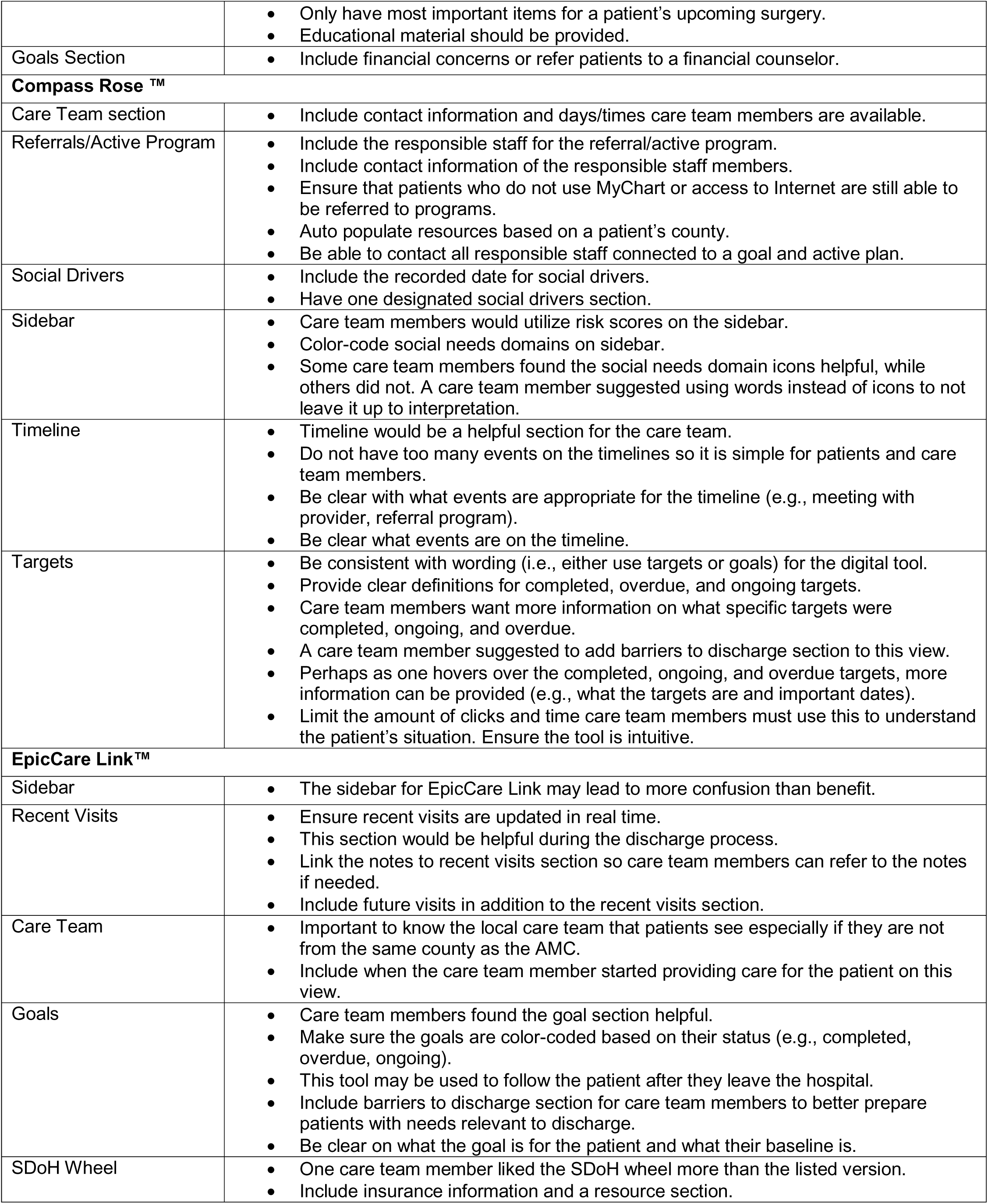
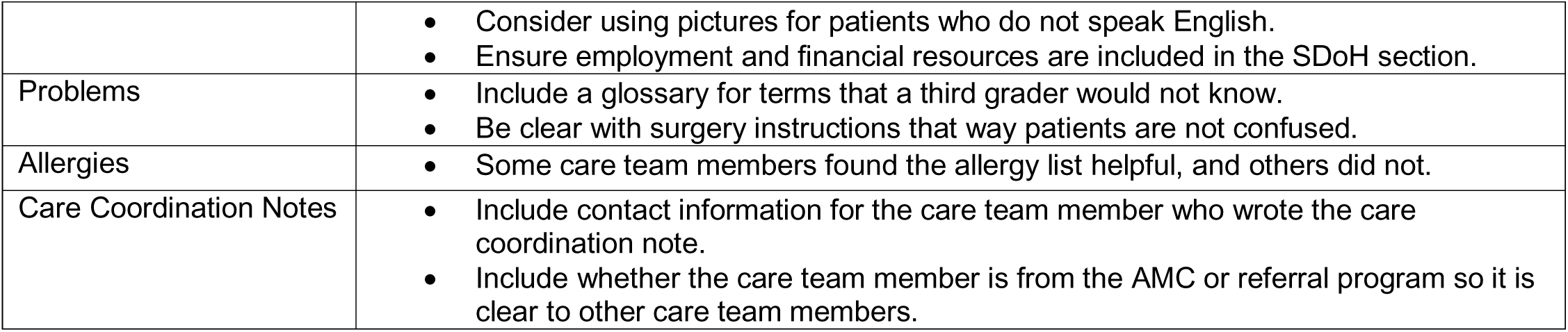
Care team usability sentiments and exemplary quotes on technology and features.

**Table 4.**
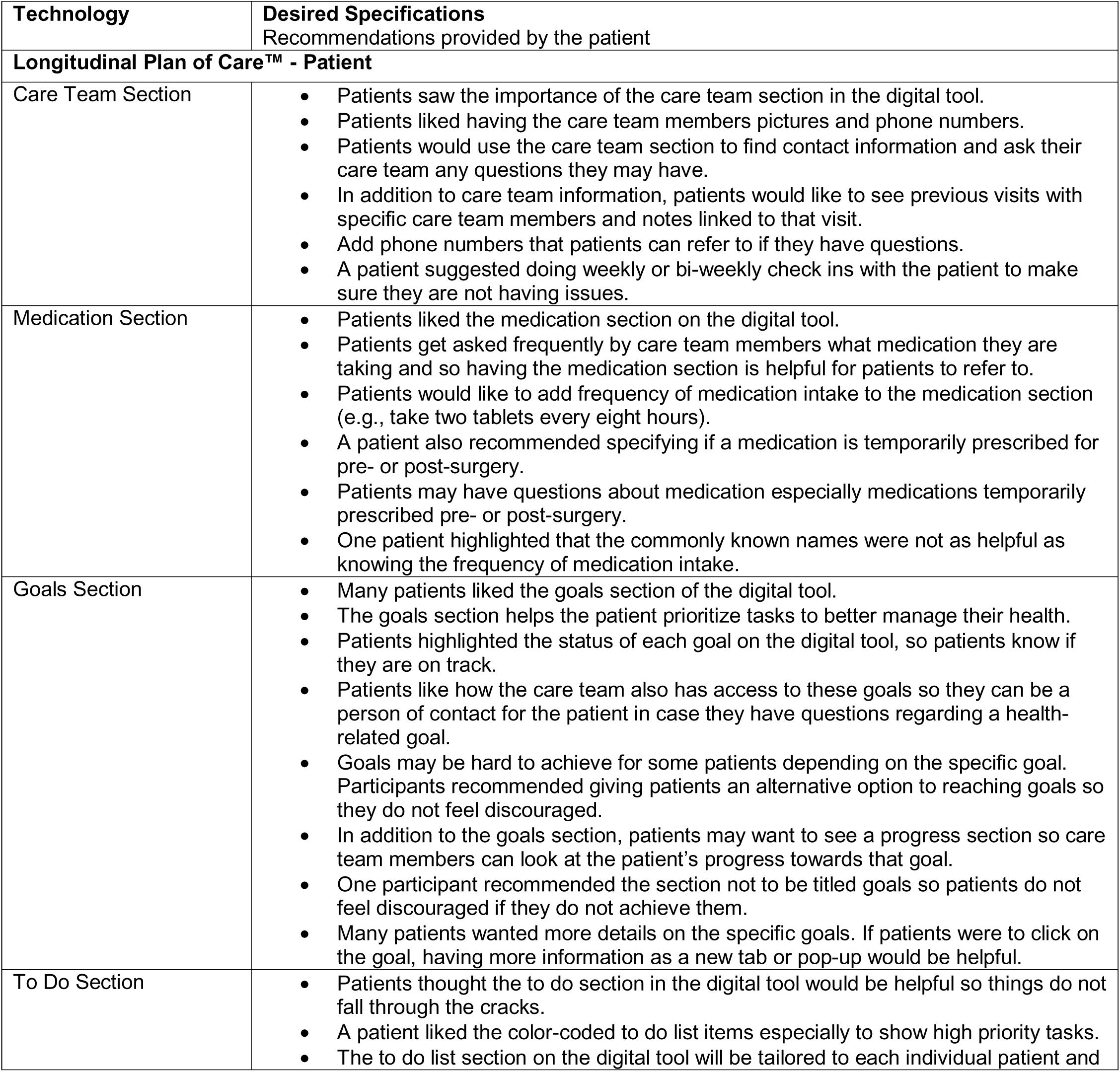
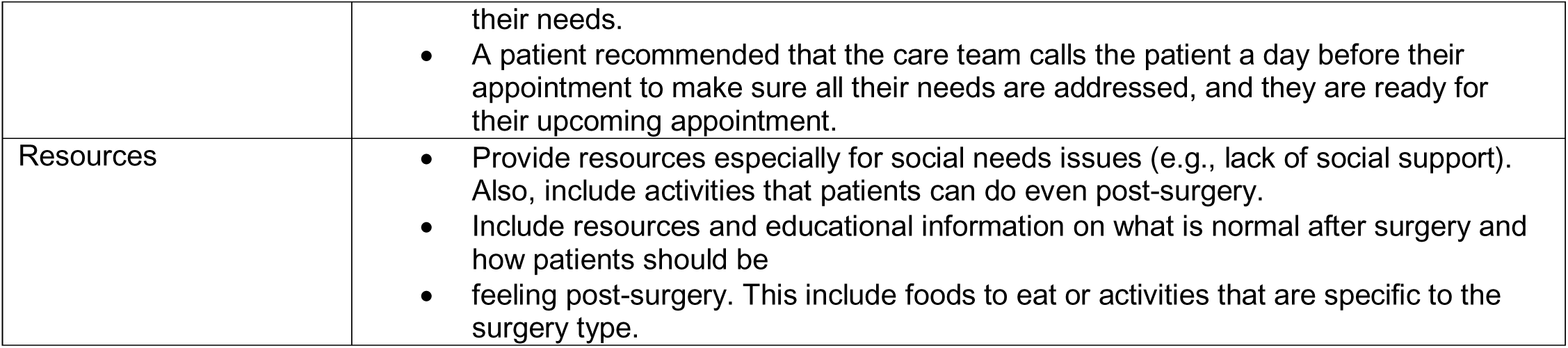
Patient usability sentiments and exemplary quotes on technology and features.

## Discussion

### Principal findings

There were several convergent themes among patients and care team members. Health-related needs collection processes varied greatly for patients and care team members, supporting the idea that there is currently no established or widely used tool to accurately identify, integrate, and address health-related needs in the perioperative period at our AMC. A few care team members had a process for screening health related needs, but most did not. Many care team members found the screening tools in the EHR (i.e., EPIC SDoH Wheel) not helpful and did not use the tool. Based on the literature, even with screening tools in the EHR, there is a variation in consistency of documentation,^45^ which aligns with our formative interviews and our chart review. There were various ways patients remembered sharing health related needs with their care team. Screening surveys and small talk were also mentioned in the formative interviews, which aligns with another previously published study.^28^

Both care team members and patients understood the benefits of the collection of health-related needs. Care team members emphasized that collection of health-related needs can better prepare patients and themselves to handle such needs. Care team members mentioned that some patients may be reluctant to report health related needs, which also aligned with our Phase I patient interview data and existing evidence.^28^ Both groups mentioned that the collection and management process need to be streamlined to improve effectiveness. Moreover, research highlights the importance of fostering trust as an important antecedent to successful patient engagement with respect to addressing health related needs.^41^ Both care team members and patients shared experiences with resolved health-related needs and unresolved needs. They also both highlighted the importance of a closed loop process when addressing health related needs. Since awareness of treatment goals has been found to improve health outcomes,^46^ integrating goals for health-related needs in a closed loop process could ensure that these patients’ needs are screened and addressed effectively.

Collection of social needs is particularly challenging. Both patients and care team members noted the same top three basic social needs. Transportation was the most frequently mentioned basic need for both groups. Both groups identified social support and home care as the top complex social need. Social support has been linked to improved health outcomes, with the most noted basic and complex social needs mentioned in our interviews aligned with the literature.^11^ At the national level, our analysis of the National Cancer Institute’s Health Information National Trends Survey (HINTS) indicated that food, transportation, and housing account for >50% of reported social needs, which align with some of the top social needs identified in our formative interviews.

There were also divergent themes between participants. Patients felt the documentation process was redundant. Patients could not recall many details regarding the collection process, or some patients lacked familiarity with the concept of health-related needs, contributing to the disconnect. Some care team members expressed that they felt uncomfortable discussing sensitive health-related needs information (i.e. finances) with patients. There were also different perspectives surrounding resources and referrals. Some patients were not aware of resources offered by their care team. Care team members expressed that the current resources are inadequate, which is a national issue given the need for multi-sectoral partnerships and capacity building on various disinvested communities.

### Design requirements and refinements needed to meet end-user preferences and needs

Based on the formative interviews, we selected four example tools that can be readily available and can be easily integrated in current workflows and the EHR. We noted variation in sentiment for all four tools in the care team design interviews. Patients also had varying levels of sentiment for each feature of the LPOC patient view. There were some features perceived as positive or slightly positive sentiment (e.g., referrals, care team information). Participants did share their suggestions to improve specific features (e.g., goals, active plans, SDoH Wheel, medications) of the tools. Given the varying levels of sentiment and participant recommendations, we determined that a customizable set of tools in Epic® or a set of tools hosted through an independent digital platform for the care team and patients were two plausible options to meet user needs. Tailored health goals and customizable features on the digital solution aligns with current practices in the literature.^47,48^

### Implications and recommendations for future design and practice

A common recommendation from the care team was to make sure that the features (e.g., care team, goals, active plans) on the digital tool included relevant information to the patient’s specific surgery. Care team participants also wanted more information provided to patients using specific features (e.g., goals and active plans). Care team members also shared that the LPOC™ patient view and CHW-facing view of EpicCare Link™ should use non-clinical terms so individuals with various backgrounds can easily understand their goals and to-do list items. Care team members also mentioned that there should be an education section to provide resources on the patient-facing digital tool. Recent visits and potentially including a future visits section on the EpicCare Link™ dashboard would be beneficial. There was discordance on the design of the SDoH section. Some care team participants liked the SDoH Wheel as a graphic, while others preferred a list or tab section to not have to interpret icons.

Patients found each section of their view of LPOC™ helpful to manage their goals and surgery journey. Patients wanted their view of LPOC™ to have more information on their care team, medications, goals, and include educational resources that pertained to their specific surgery (e.g., recommended diet post-surgery). One patient felt that the goals section needed to be reworded and should provide goal alternatives, so patients feel motivated to achieve their health goals. In future iterations of this work, we plan to deploy prototypes of our digital solution tools and conduct usability sessions with care team members and patients. A final solution, whether through Epic® or an independent platform, will need additional workflow integration considerations.

### Limitations

The current study had several limitations. First, our study was not linked to an evaluation of specific health outcomes.^27^ Our formative interviews were conducted at outpatient clinics associated with our AMC; albeit our AMC is one of the largest health systems in the Midwest and delivers complex surgery for a diverse pool of patients who live in the AMC city and across the state. We did, however, conduct convenience sampling for participants in our study, which may have impacted our formative interviews. In addition, some patients may not have fully understood what health related needs were during interviews which could have impacted our results; however, our team followed systematic procedures to ensure that patients were made aware of the concepts covered in our interview sessions and we documented challenges around awareness as well.

## Conclusion

In the current study, we obtained qualitative feedback from both care team members and patients in our AMC’s Department of Surgery on how to collect and manage health-related needs, including social needs. We conducted thematic analysis to identify convergent and divergent themes about critical gaps and potential solutions. Health-related needs collection and resolution processes varied greatly between the perspectives for both care team members and patients. Both patients and care team members frequently mentioned social support and home care as their top social need. Based on the formative interviews, we selected digital tools from Epic® as potential digital solutions to present to a subset of our participants. We identified a desired set of features in an ideal digital solution based on our sentiment analysis of the design sessions. Further research is necessary to continue improving our proposed solution to promote health-related needs screening and management for complex surgery patients.

## Data Availability

Data is confidential and cannot be shared to respect privacy of participants.

## Other Acknowledgments

We are grateful to the patients and care team members who participated in this study.

## Author Contributions

All authors contributed to the design, writing, and review of the manuscript. Dr. Fareed led in the writing of the manuscript. Drs. Odysseas, Tsilimigras, and Pawlik, along with Humza Asgher, Giovanni Catalano, Bridget Hartwell, and Kathleen Bolton contributed to the original draft. Formal analysis was led by Dr. Fareed and Humza Asgher. All authors provided review and editing for subsequent versions of the manuscript.

## Abbreviations

AMC: Academic Medical Center
APP: Advanced Practice Provider
CBO: Community-Based Organization
CR: Compass Rose
EHR: Electronic Health Record
GST: Goal-Setting Theory
LPOC: Longitudinal Plan of Care
OOI: Ohio Opportunity Index
PCRM: Patient Care Resource Manager
SCT: Social Cognitive Theory
SDoH: Social Determinants of Health
SMART: Specific, Measurable, Achievable, Relevant, Time-bound

