## Supplementary material for "Exploring a digital health solution to collect and manage health-related needs for complex surgery patients: Mixed-methods study": S1 Patient Interview Guides.pdf

### Supplementary File 1

#### Supplement 1. Patient Interview Guide Phase I

**RISE: Reporting and Addressing Health-related Needs Information to Improve Surgical Outcomes and Experience**

**Needs Assessment Interview Guide – Patient**

**BACKGROUND**

First, let us THANK YOU for agreeing to participate in our research project. We are researchers from The Ohio State University. We are contacting you as part of a project we are doing that pertains to the development and use of a technology-based smartphone application for surgery patients. **For today's call, we are particularly interested in your experiences and preferences with reporting, collecting, and managing non-medical social needs information before, during, and after surgery at Ohio State.** Examples of non-medical social needs information include home environment, safety, discrimination, access to food, childcare, transportation, and financial security. Results from the study will help us identify gaps in the current practices of care and guide the development of a user-centered smartphone application to improve surgical outcomes. We have scheduled the next 40-60 mins to discuss these topics.

**INTERVIEW TOPICS**

In this interview, we will ask you a series of open-ended questions to get your perspectives about different topics related to the workflow for surgical care. As an overview, these topics are:

Section 1: Background - understanding you and your health care experience
Section 2: Clinical care
Section 3: Information about non-medical social needs
Section 4: Desired features for reporting non-medical social needs

**Section 1: Background**

- How old are you?
- What is your race?
- Are you Hispanic or Latinx?
- In what county do you live?
- **[For patients who will undergo surgery].** For our records, can you state the surgical procedure you plan to have at Ohio State and when it is planned?
- **[For patients who underwent surgery].** For our records, can you state the surgical procedure you received at Ohio State and when it was completed?

**Section 2: Clinical care**

1. Can you tell us about your experience with the care you have received so far?
a. How do you communicate with your care team?
    b. Who are the care team members that help you with your care and how do they help you? **[Clarify if needed: who do you talk to the most? what do you typically talk about? how does your doctor, nurse. or case manager help you?]**
    c. Have you experienced any challenges in following your care team recommendations?

<u>**Section 3: Information about non-medical social needs**</u>

We want to know about non-medical factors that affect a person's health before, during, and after surgery. **[Make sure to list: Examples of these social and environmental factors include a person's: home environment (mold, air quality, clean water, safe living space), family and social support, fair treatment (no discrimination), and access to quality food and transportation)]**

1. Have you ever shared non-medical social needs information with your care team member?

**If yes,**

- Who specifically? Case manager? Nurse? Physician?
- Did they do anything about it?
- How did you feel about this experience?
- How did you share this information? Verbal? Phone? Patient Portal Message?
- Did you ever have a referral made to a social need resource (e.g., food pantry, taxi service)?
- [for Oncology patients] Have you reported any needs using the distress survey? How many times do you recall filling this survey? What do you think about this survey?

**If no,**

- What would make you comfortable enough to share this type of information with your care team member?

**General,**

- What are the top social need priorities you think care team members should always focus on?
- Do you have any general suggestions about how non-medical social needs information and issues can be handled by your care team?

<u>**Section 4: Desired features for reporting non-medical social needs**</u>

We are creating a smartphone application for reporting social needs to your care team. You can communicate any non-medical social needs to a care team member before, during, and after your surgery. Someone from your care team can use this information to connect you to resources in your neighborhood to help you overcome challenges you may face with managing your surgery and overall health.

1. Do you have a smartphone?
2. What do you primarily use the smartphone for?
3. Do you access the Ohio State MyChart application with it?

4. Do you have any challenges with using applications such as MyChart on your smartphone?
5. Is there anything you would like to see in a smartphone app to help you report non-medical social needs?
6. Would you use an application to report non-medical social needs on your smartphone?
    - What information would you like collected through this app?
    - What are your preferences for collecting this information (e.g., before your clinic visit? Continuously?
7. What additional training do you anticipate with using such an application?
8. How would you prefer to learn to use this application? (e.g., videos, flyers, in-person training)?

**INTERVIEW CLOSURE AND FOLLOW-UP**

- Is there anything else we should know or that you would like to tell us about sharing non-medical information before, during, or after surgery?

- Would you be interested in being contacted in the future to participate in a group to help explore the topics we covered in this interview?

**THANK YOU so much for your time and participation. Your comments and this discussion were extremely helpful.**

### Supplement 2. Patient Interview Guide Phase II

**RISE: Reporting and Addressing Health-related Needs Information to Improve Surgical Outcomes and Experience**

**User Experience Guide – Patient**

**Moderator checklist:**

- Have Miro open and set to Frame 6 –
- Teams open to chat with second interviewer
- Script open

**[Interviewer will take notes on Miro on their laptop]**

**Background**

First, let us THANK YOU for joining us for the follow-up interview. Based on what we discovered from the first phase of our study, we would like to talk to you about how we can improve patient activation among surgery patients at The Ohio State University. Increasing patient activation levels can lead to better surgery outcomes and reduce patient readmissions and surgical complications. **We define patient activation as improving a person's confidence, self-management skills (i.e., controlling your behavior and actions), and self-efficacy, which is believing in yourself and your ability to prepare for surgery and recovery.** This can be achieved by addressing 1) patient health-related social needs (HRSNs) (e.g., reliable housing, transportation); 2) healthcare needs (e.g., high blood pressure, diabetes) and 3) health behaviors like diet, alcohol and tobacco use.

Our purpose today is to obtain your feedback on how a digital tool can support these needs and improve patient activation. We have scheduled the next 40-60 mins to discuss these topics.

**Introduction to the Interview**

Patients will complete assessments that focus on social needs, healthcare needs, and health behaviors. The care team will work with patients to set and achieve goals in these areas. A community health worker will help with addressing social needs goals, and case managers and social workers will coach patients on social needs goals. A nurse practitioner will help patients with health behavior and healthcare goals. A patient navigator will manage the assessment and referral processes, and make sure patients achieve their goals.

To be explicit, we are testing the design of a summary tab that would appear in MyChart that provides information on goals focused on health-related social needs, healthcare needs, and health behavior needs. I'm going to ask questions on a potential design to help us prioritize features in the digital solution. It will be helpful if you can provide as much information as possible about how you're processing the digital solution. As you answer questions, if you are looking for information, are confused by anything, or like or dislike something, please vocalize these normally silent thoughts. There are no right or wrong answers to the questions I'm going to ask, and the information you provide will give critical information to the design team.

Do you have any questions so far?

**Show Example of SMART goal in action plan (Frame 6)**

This is an example of an action plan for a surgery patient. It is organized as a SMART goal which stands for: **specific, measurable, attainable, relevant, and time-bound** to help improve patient activation. In this example, the patient needs to make sure they have a high-protein diet after their surgery to have a smooth recovery. This health behavior SMART goal serves as an educational tool, outlining what is required to have a successful surgery.

**Show Patient view of Longitudinal Plan of Care (Frame 7)**

**Have you seen this before?**

**This is the patient view in MyChart**

This is what you or your caregiver would view in the MyChart application. Goals based on responses would be displayed on the top left. Below goals we have a to do list with items linked to the action plan which are the steps the patient and care team would complete to achieve the goals. On the top right is a list of medications a patient may be on, and below is the care team's information.

[Give them 15 seconds to look at prototype]

**Questions:**

b. Given what you understand with what we're trying to do, what do you think about this tool?
c. Do you think this would help address SMART goals patients may have?
    a. If not, how would you refine this tool to help patients achieve their SMART goals?
d. How should high priority goals (HRSNs, healthcare needs, and health behavior) be displayed on the dashboard?

**Yes/No exercise:**

For each digital solution, we would like you to share which components should be included to improve patient activation levels.

- [If participants are not sure which features go in a category: drive the conversation and specifically ask them about a component one by one – from left to right]

[Second interviewer will create bins for participants into the categories they tell you to]

**Follow-up questions**

e) Overall, what are your thoughts about potentially using this digital solution?
e) If you could change two things about the digital solution, what would they be?
e) If we redesign this digital solution based on those recommendations, is there anything you think we *shouldn't* change?
e) (If applicable) During the interview, I noticed you did/said ____. Could you tell me more about that?

e) Anything else?

Thank you again for your time. Your feedback has been very helpful. Have a good day!
