## Supplementary material for "Exploring a digital health solution to collect and manage health-related needs for complex surgery patients: Mixed-methods study": S2 Care Team Interview Guides.pdf

### Supplementary File 2

#### Supplement 1. Care team Interview Guide Phase II

**RISE: Reporting and Addressing Health-related Needs Information to Improve Surgical Outcomes and Experience**

**Needs Assessment Interview Guide – Care Team**

**BACKGROUND**

First, let us THANK YOU for agreeing to participate in our research project. We are researchers from The Ohio State University. We are contacting you as part of a project we are doing that pertains to the development and use of technology-based smartphone and web portal applications for surgery patients. **For today's call, we are particularly interested in individuals' experiences and preferences with reporting, collecting, and managing non-medical social needs information before, during, and after surgery at Ohio State.** Examples of non-medical social needs information include home environment, safety, discrimination, access to food, childcare, and transportation, and financial security. We are interested in establishing how you provide care during the treatment trajectory for your patients. Results from the study will help us identify gaps in the current practices of care and guide the development of user-centered smartphone and portal applications to improve surgical outcomes. We have scheduled the next 40-60 mins to discuss these topics.

**INTERVIEW TOPICS**

In this interview, we will ask you a series of open-ended questions to get your perspectives about different topics related to the workflow for surgical care. As an overview, these topics are:

Section 1: Background - understanding your role and care workflow
Section 2: Clinical care
Section 3: Information about non-medical social needs
Section 4: Desired features for managing non-medical social needs

**Section 1: Background**

- How old are you?
- What is your race?
- Are you Hispanic or Latinx?
- What is your current role and how long have you been in that role?
- In which surgery department(s) do you provide care?

**Section 2: Clinical care**

1. Please describe the typical course of treatment for a new patient who undergoes surgery during the pre-op, peri-op, and post-op stages. **[Clarify if needed: what is the treatment trajectory for your patients? When and how does your role impact this trajectory?]**

- Please describe the process of engaging with a patient from their first pre-op encounter through to their post-surgery discharge
- What are the challenges with adherence among your patient population?
- Can you characterize the patients that you manage?
- How do you typically communicate with them?

**Section 3: Information about non-medical social needs**

We want to know about non-medical factors that affect a person's health before, during, and after surgery. Examples of these social and environmental factors include a person's: home environment, family and social life, fair treatment (no discrimination), and access to quality food and transportation, and minimal to no exposure to poor air quality (including smoking).

1. Do your patients report non-medical social needs information?

**If yes,**

- What prompts and tools do you use to collect this information?
- What specific information is recorded and where is it recorded?
- Who has access to this information in your care team?
- What do you do with this information?
- How do you think this information can affect patient care before, during, and after surgery?
- Have you ever made referrals to resources (e.g., food pantry, taxi service, medication assistance) based on information about social needs?
  - What is that process like?
  - Have you or the patient faced challenges with this process?

**If no,**

- What can be an ideal approach to collecting this type of information?
- Do you see any value to collecting and using this type of information for patients and the care team?

**General, [Clarify if needed: is the current collection approach effective?]**

- Do you have any general suggestions about how non-medical social needs information and issues can be handled by your care team?
- **What are the top social need priorities you think care team members should always focus on?**

**Section 4: Desired features for managing non-medical social needs**

We are creating smartphone and portal applications for reporting social needs to your care team. Patients can communicate any non-medical social needs to a care team member before, during, and after your surgery, and this information will be displayed via a dashboard on the portal. Someone from your care team can use this information to connect a patient to resources in their neighborhood to help them overcome challenges they may face with managing their surgery and overall health.

1. Have you ever used the EPIC electronic health record system to screen and view information about a patient's non-medical social needs? **[Clarify if needed: do you use in-basket messages, notes, a flow sheet, or the EPIC SDoH wheel to record and manage social needs]**
**If yes,** what do you think about how this information is presented?
**If no,** why not?
2. How would you like to see information about patient social needs in the EPIC electronic health record system?
    - Are there particular features (information type and functions) of a portal that would be helpful?
    - Do you think patients can use a smartphone app to report this information?
3. What additional training do you anticipate with using such an application?
4. How would you prefer to learn to use this application? (e.g., videos, flyers, in-person training)?
5. Are there any other suggestions you have for this portal?

**<u>INTERVIEW CLOSURE AND FOLLOW-UP</u>**

- Is there anything else we should know or that you would like to tell us about sharing non-medical information before, during, or after surgery?

- Would you be interested in being contacted in the future to participate in a group to help explore the topics we covered in this interview?

**THANK YOU so much for your time and participation. Your comments and this discussion were extremely helpful.**

### Supplement 2. Care team Interview Guide Phase II

**RISE: Reporting and Addressing Health-related Needs Information to Improve Surgical Outcomes and Experience**

**User Experience Guide – Care Team**

**Moderator checklist:**

- Have Miro open and set to Frame 1
- Teams open to chat with second interviewer
- Script open

**Background**

THANK YOU for joining us for this follow-up interview. Based on what we discovered from the first phase of our study, we would like to talk to you about how we can improve patient activation among surgery patients at The Ohio State University. Increasing patient activation levels can lead to better surgery outcomes and reduce patient readmissions and surgical complications. **We define patient activation is defined as improving a person's confidence, self-management skills, and self-efficacy in preparation for their surgery and recovery.** This can be achieved by addressing 1) patient health-related social needs (HRSNs) (e.g., reliable housing, transportation); 2) healthcare needs (e.g., hypertension, diabetes) and 3) health behaviors like diet, alcohol and tobacco use.

Our purpose today is to obtain your feedback on how specific digital tools can support these needs and improve patient activation, and how they can be integrated into the workflow. We have scheduled the next 40-60 mins to discuss these topics.

**Introduction to the Interview**

Patients will complete assessments that focus on the three main contributors to patient activation (HRSNs, healthcare needs, and health behaviors). Care team members will work with patients to develop goals around these three areas. Based on these goals, a community health worker will assist with addressing HRSN goals, and case managers and social workers will coach patients on HRSNs goals. An Advanced Practice Provider (i.e., nurse practitioner) will guide patients on health behavior and healthcare goals. A patient navigator will oversee the assessment, referral, and intervention processes, stepping in as needed to ensure patients achieve their goals.

During our call, I'm going to ask questions on potential designs and guide you through a card-sorting exercise to help us prioritize features in the digital solution. It will be helpful if you can provide as much information as possible about how you're processing the digital solution. As you answer questions, if you are looking for information, are confused by anything, or like or dislike something, please vocalize these normally silent thoughts. There are no right or wrong answers to the questions I'm going to ask, and the information you provide will give critical information to the design team.

Do you have any questions so far?

**The main interviewer shares their screen, where Miro is pulled up in presentation mode (Frames 1-5 in Miro).**

**[Interviewer 2 should also have Miro open and ready for the card-sorting activities]**

Thank you. I'm going to start the recording. I'm going to bring up the prototype on my computer, and then I'll start sharing my screen with you.

**Example of SMART goal in action plan (Frame 1)**

This is an example of an action plan for a patient. It is organized as a SMART goal which stands for goals that are: **specific, measurable, attainable, relevant, and time-bound** to help improve patient activation. In this example, the patient needs to make sure they have a high-protein diet after their surgery to have a smooth recovery. This health behavior SMART goal serves as an educational tool, outlining what is required to have a successful surgery.

***If participant is clinician (APP, physician, resident): show LPOC and CR. If extra time, show Care Link**

***If participant is CHW, social worker, or case manager: show CR and Care Link. If extra time, show LPOC.**

**Longitudinal Plan of Care Dashboard – care team view (Frame 2)**

The Longitudinal Plan of Care (LPOC) is an Epic-based program that provides a detailed overview of a patient's treatment plan using patient goals and active plans. LPOC focuses on the patient's current diagnoses, concerns, and potential health risk factors.

This is a customizable digital tool [**describe the dashboard from left to right** (e.g., care coordination notes, allergy and problem lists are on the left. Social Determinants of Health (SDoH) wheel and goals are in the middle and active plans on the right side)]. The SMART goals I introduced in the previous slide would be listed under the Goals section. The active plans on the top right would be the interventions in place (achievable/attainable part of the SMART framework) to help the patient achieve their HRSN, health behavior, and healthcare goals.

- e.g., imagine a patient has missed doctor appointments due to lack of transportation. In the active plan section, the care team would see that the patient was referred to a local transportation service.

[Give them 15 seconds to look at prototype]

**Questions:**

a. Given what you understand with what we're trying to do, what do you think about this tool?
b. Do you think this would help address SMART goals patients may have to achieve better patient activation?
    a. If not, how would you refine this tool to help patients achieve their SMART goals related to surgical patient activation?

[If someone asks you if we use this tool: Say that it varies by department, and we are showing you what is possible from the vendor side.]

**Card-Sorting exercise:**

For each digital solution, we would like you to rank the seven components into four categories (least important, moderately important, most important, and other)?

- [If participant asks what we mean by importance: In terms of value for you, will you use it in order to move the needle in the three drivers of patient activation (HRSNs, health behavior, and healthcare needs)?]

- [If participants are not sure which features go in a category: drive the conversation and specifically ask them about a component one by one – from left to right]

[Second interviewer will create bins for participants into the categories they tell you to]

**Longitudinal Plan of Care – Patient View (Frame 3)**

This is the patient's point of view for Longitudinal Plan of Care. Goals based on responses would be displayed on the top left for the patient. Below goals we have a to do list with items linked to their active plan.

[Give them 15 seconds to look at prototype]

**Questions:**

a. Based on what you are seeing, do you think this would improve a patient's activation level?
    a. If so, how and why?
    b. If not, how would you refine this tool to help patients improve surgical patient activation levels?

***No card sorting activity for care team members on patient view of LPOC***

**Show CR Dashboard – care team view only (Frame 4)**

Compass Rose (CR) is an Epic-based program that can be used to refer patients with health-related social needs and health behavior needs to community services (e.g., nutrition assistance, group homes). CR allows you to identify resources for the patient and communicate with the local resources in the patient's community. CR allows you to embed and track all the necessary steps (e.g., engagement with a CHW, completing application paperwork) to get the patient the resources identified for them and get their needs addressed. CR displays the status and documents any issues with each step.

- [**Describe the dashboard from left to right** (e.g., Targets are the goals for the patients with the timeline related to these steps below. On the right, we see the social drivers of health addressed.)]

[Give them 15 seconds to look at prototype]

**Questions:**

a. Based on what you see, how do you think this would improve a patient's activation level?
b. Are there any refinements you would make to this dashboard to improve a patient's activation level?

**Card-Sorting exercise:**

[Second interviewer will create bins for participants into the categories they tell you to]

**For case managers, social workers, and community health workers or if participant has extra time:**

**Show Care Link Dashboard (Frame 5)**

With EpicCare Link and Healthy Planet Link, community-based organizations (CBOs) can access care management tools such as message the care team at OSUMC, add to the longitudinal plan of care, and resolve patient care gaps. Community health workers (CHWs) would be able update care coordination notes, goals, and social needs in the EHR.

- [**Describe the dashboard from left to right** (e.g., Care coordination notes, allergy and problem lists are on the left. Social Determinants of Health information and goals are in the middle. Information on care team and recent visits are on the right.)]

[Give them 15 seconds to look at prototype]

**Questions:**

a. Based on what you see, how do you think this would improve a patient's activation level?
b. Are there any refinements you would make to this dashboard to improve a patient's activation level?

Probe: How could we incorporate notes or updates from Community Health Workers (CHWs) or other care team members to increase patient activation?

**Card-Sorting exercise**

[Second interviewer will create bins for participants into the categories they tell you to]

**Final questions:**

a. Do you think it also helps to collect vitals and put it in one of these tools (pulse, temperature, blood pressure, respiratory rate, oxygen saturation levels)?
b. Would you prefer LPOC or CR or a combination or something else to improve surgical patient activation levels?
c. Do you have any questions for us?

Thank you again for your time. Your feedback has been very helpful. Have a good day!
