## Supplementary material for "Exploring a digital health solution to collect and manage health-related needs for complex surgery patients: Mixed-methods study": S3 COREQ guidelines.pdf

### Supplementary File 4

#### Summary of COREQ guidelines and study activities

| No. Item | Guide questions/description | Reported |
|---|---|---|
| **Domain 1: Research team and reflexivity** | | |
| *Personal Characteristics* | | |
| 1. Interviewer/facilitator | Which author/s conducted the interview or focus group? | HA, GC, OC, NF |
| 2. Credentials | What were the researcher's credentials? e.g. PhD, MD | HA, BS; DT, MD, PhD; GC, MD; OD, MD; KB, MS, RN, AGCNS-BC, OCN; BH, MSN, APRN-CNP; TP, MD, PhD, MPH; NF, PhD, MBA |
| 3. Occupation | What was their occupation at the time of the study? | OSUMC Research Staff |
| 4. Gender | Was the researcher male or female? | KB, BH - female<br>HA, DT, GC, OC, TP, NF - male |
| 5. Experience and training | What experience or training did the researcher have? | Diverse range of qualitative interviewing experience, but all research team members were trained prior to conducting interviews. All staff had experience with our RISE study. |
| *Relationship with participants* | | |
| 6. Relationship established | Was a relationship established prior to study commencement? | Research team members who work at the AMC may have interacted with participants prior to the study. Research team members who conducted interviews did not know participants prior to study commencement. |
| 7. Participant knowledge of the interviewer | What did the participants know about the researcher? e.g. personal goals, reasons for doing the research | All participants were briefed on the objectives of the study during the screening, introduction, and consent processes before interviews. |
| 8. Interviewer characteristics | What characteristics were reported about the inter viewer/facilitator? e.g. Bias, assumptions, reasons and interests in the research topic | DT, GC, OC, KB, BH, TP, NF had roles in developing prototypes and worked closely in the design process. |
| **Domain 2: study design** | | |
| *Theoretical framework* | | |

| | | |
|---|---|---|
| 9. Methodological orientation and Theory | What methodological orientation was stated to underpin the study? e.g. grounded theory, discourse analysis, ethnography, phenomenology, content analysis | Thematic analysis |
| *Participant selection* | | |
| 10. Sampling | How were participants selected? e.g. purposive, convenience, consecutive, snowball | Convenience sample for both care team members and patients. |
| 11. Method of approach | How were participants approached? e.g. face-to-face, telephone, mail, email | Participants were contacted by email or phone by the research team for recruitment. |
| 12. Sample size | How many participants were in the study? | 44 (20 care team members and 24 patients) |
| 13. Non-participation | How many people refused to participate or dropped out? Reasons? | Phase I:<br>Eight care team members out of twenty-eight (29%) could not participate due to scheduling conflicts or non-availability. Twenty-seven patients out of fifty-one (53%) did not participate because they either refused or did not show up to the interview after accepting our invitation.<br>Phase II: Three care team members could not participate due to scheduling conflicts or non-availability. Fourteen patients out of 19 (70%) either refused or did not show up to the interview after accepting our invitation. |
| *Setting* | | |
| 14. Setting of data collection | Where was the data collected? e.g. home, clinic, workplace | Virtually (Zoom) for care team members. There was a mix of virtual (Zoom) and in-person interviews at one of the AMC's locations. |
| 15. Presence of non-participants | Was anyone else present besides the participants and researchers? | It is not clear if others were present off camera during virtual interviews. Some patients may have had family or friends present during in-person interviews. |
| 16. Description of sample | What are the important characteristics of the sample? e.g. demographic data, date | Care team member role, OOI scores of patients' residential communities, racioethnicity of patients |
| *Data collection* | | |
| 17. Interview guide | Were questions, prompts, guides provided by the authors? Was it pilot tested? | Interview guide was designed by our senior researcher (NF). Pilot interviews were conducted for care team members and mock patients. |
| 18. Repeat interviews | Were repeat interviews carried out? If yes, how many? | No |
| 19. Audio/visual recording | Did the research use audio or visual recording to collect the data? | Audio recording of Zoom and in-person interview which were transcribed using Zoom. |

| | | |
|---|---|---|
| 20. Field notes | Were field notes made during and/or after the interview or focus group? | Interviewers may have taken notes during and/or after the interview, but these were not included in our data. |
| 21. Duration | What was the duration of the interviews or focus group? | Approximately 60 minutes. |
| 22. Data saturation | Was data saturation discussed? | Interviews and the research team met once a week to discuss emerging themes to ascertain saturation, including discussions around recurrence of specific themes. |
| 23. Transcripts returned | Were transcripts returned to participants for comment and/or correction? | No |
| **Domain 3: analysis and findings** | | |
| *Data analysis* | | |
| 24. Number of data coders | How many data coders coded the data? | Two coders and one senior researcher. |
| 25. Description of the coding tree | Did authors provide a description of the coding tree? | NF and HA developed a coding tree and discussed this with the rest of the research team. |
| 26. Derivation of themes | Were themes identified in advance or derived from the data? | Parent codes were based on the semi-structured interview guides and child codes were derived from our formative interviews. |
| 27. Software | What software, if applicable, was used to manage the data? | Microsoft Word |
| 28. Participant checking | Did participants provide feedback on the findings? | No |
| *Reporting* | | |
| 29. Quotations presented | Were participant quotations presented to illustrate the themes/findings? Was each quotation identified? e.g. participant number | Yes, see Results and Appendix for participant quotations. At least three quotations were found for each child code in our tables. Care team members were identified by role. Patients were identified by age, gender, surgery type, racioethnicity and OOI group. |
| 30. Data and findings consistent | Was there consistency between the data presented and the findings? | Yes, and this is described in the discussion section. |
| 31. Clarity of major themes | Were major themes clearly presented in the findings? | Major themes are presented in the Results and further analyzed in the discussion. |
| 32. Clarity of minor themes | Is there a description of diverse cases or discussion of minor themes? | Discussion of minor themes are included in the paper. |
