## Supplementary material for "Exploring a digital health solution to collect and manage health-related needs for complex surgery patients: Mixed-methods study": S4 Patient Activation View.pdf

### Supplementary File 5

Example SMART goal that would align with their surgery available through MyChart and as a print out with the AVS:

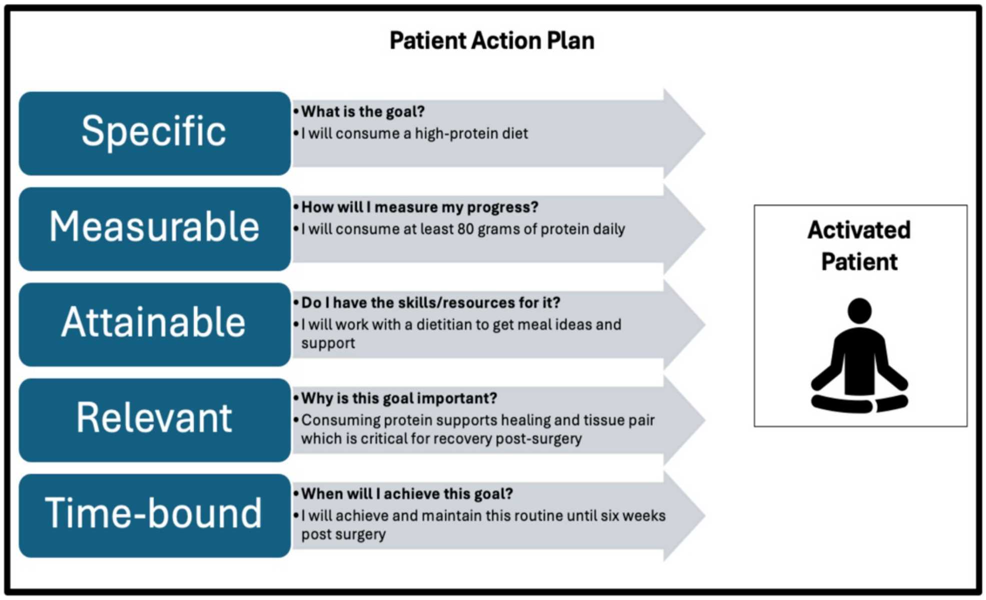
