## Supplementary material for "Exploring a digital health solution to collect and manage health-related needs for complex surgery patients: Mixed-methods study": S5 Current Needs Tables.pdf

### Supplementary File 6

#### Supplemental Table 1. Current medical care needs and engagement

| Theme | Care Team Quotes | Patient Quotes |
|---|---|---|
| **Engagement:** How care team and patient seeking clinical care interact | Case Manager 1: “like 50 to 75 [Percent] is on the phone. A lot of our patient outreach is done by phone but we do try to see all of our patients in person when they're in clinic. So we do a portion of patient contact in person as well. But it really depends. You know how often they're in clinic, and when they're coming in.”<br><br>Physician 7: “Yeah, so we have 2 types of patients. One are elective, and one are like kind of urgent in hospitals. So for the elective patients, they'll typically see me in clinic. I will then you know, go through anything. If there's any additional testing they need. They'll get that testing done and then come back and see me. And then we'll schedule surgery. Surgery will happen. They'll be in the hospital, for you know. on average 5 to 7 days. Go home or to rehab facility, if that's what they need. And then they'll follow up with me and usually like 4 to 6 weeks for urgent inpatient. So usually it's the same. Operative and post operative course, it's just the beginning part the workup is done in the hospital, and that's where I meet them. And then, would you say, your interactions with patients are mainly in person during their pre app and post OP. Visits, and obviously surgery or, yes.”<br><br>Advanced Practice Provider 3: “I see the patient first. I do a thorough history and physical examination and then we meet myself and the surgeon, and and discuss the patient in the case, and review imaging, and then the surgeon goes and evaluates the patient. We then, you know, decisions are made between the patient, the surgeon, and you know myself as applicable in terms of what the plan will actually be. If surgery is what the plan is, we will either set up another pre up appointment to consent and go over things. It depends on the timing of | Patient 11 (male, in their 70s, gastrointestinal/benign, NH white, most opportunity): “The doctor came in every morning, explained what was going on and kept me updated. And finally they decided they had to operate. So that's what we went from there. And that's about all I know My daily meeting with the doctors. They would come in in the morning and tell me what was going on. and the process that they were going to do.”<br><br>Patient 7 (male, in their 60s, gastrointestinal/oncology, NH white, lesser opportunity): “Well, this doctor saved my life. Basically. It's as far as the rotation of nurses and patient care technicians and the different doctors that I saw is it all on the same plane, was all on the same level. I got the same attention and the same care. and all of those are across the board and I liked that. It was very helpful for me to understand what was going on, and to step forward, because that's what I wanted to do. I wanted a positive outcome. I'm gonna fight this. And everybody. Was it on my side, you know. “<br><br>Patient 5 (female, in their 30s, gastrointestinal/benign, Hispanic, lesser opportunity): “Well, I don't really remember much of the morning like when I went in to get the procedure done, but the nurse the nurses that I had the following couple of days. Were great. They listened. I think I was on like for the fifth floor. I don't remember what room but they were very attentive, very fine. And understanding took my like complaints as far as like pain |

| | | |
|---|---|---|
| | things that the patients ready for surgery now. or if they need more chemotherapy, or if they need any chemotherapy. Perhaps they haven't started they need other referrals prior to surgery. But we always have some sort of pre-OP app appointment where we can send and do education. We normally send the patient to our opac, our pre surgery, outpatient pre anesthesia center at the James, and have them evaluated prior to surgery and then they go into surgery. and then they'll have you know it's either an outpatient ambulatory surgery, or they'll have a stay at the James for a certain amount of time for their immediate recovery from surgery, and then they go home, or to sometimes that rehab facility, and we'll see them usually 2 weeks after, in clinic for a post out visit, and then, you know, several visits after that until they're fully recovered, and then surveillance visits as appropriate.” | management seriously. they tried to encourage me to, you know, eat or drink, and they checked in regularly, just to make sure that I was okay. And if there is anything that needed to be addressed with. like the surgical team, for any reason. They would pass the message along to the appropriate person, so I received really good care. During my stay. I don’t really remember anything negative.” |
| **Patient Care:** Typical course of treatment or the patient’s clinical experience | Case Manager 1: “For about 75% of my patients, that would be upon discharge from the hospital after their surgery. I'll get added to the care team to follow needs after they've had surgery. I will get hand off from the inpatient PCRMs but for other patients sometimes the clinic nurse or nurse practitioner, one of the doctors will bring a concern to me that the patient would need something or occasionally like if I'll see that a patient is calling in multiple times with like, multiple clinical needs. Sometimes I'll pick up folks that way. With patients that just kind of have high needs and are calling in a lot. If I see people like without insurance or things like that, like on our clinic list. I can get pick up folks that way.”<br><br>Physician 2: “I typically meet the patient as a consultation, whether it's in the hospital or in my clinic. You know, assess them for surgery, assist their readiness. Most of my practice is actually at the [BLINDED] Hospital, so I see a lot of folks from disadvantaged populations to social work, especially if they're inpatient or often engaged. Outpatient I will often lean on one of the piece outpatient PCRMS through Martha Morehouse to help. If there's | Patient 8 (male, in their 30s, gastrointestinal/benign, NH white, lesser opportunity): “Yeah. So are was overall. Very good I think the team of doctors were. I think, yeah. competent. at the very least. There was some times where communication was not as good as I hoped for…I think there was just times where I was left feeling kind of in the dark about what the plan was moving forward. So, for instance, like We're not going to, for instance, take your NG tube out today, but kind of being left with not much of an idea of the plan moving forward of like. I was like but if this thing were to happen then, maybe tomorrow, or like, you're not going to be discharged today. But these are what we're looking for looking at in terms of when to, you know, discharge you just a few times like that where I was left feeling a little bit kinda in the dark or frustrated.” |

| | |
|---|---|
| | somebody who needs some additional, you know, support and they've been able to help me, even though they're patients who are at [BLINDED]. So I try to get patients pre-OP optimize as much as possible. So main things that I always try to focus on our nutrition and access to, you know, protein sources and then, smoking cessation. And you know, drug and alcohol cessation. So those are kind of the main ones that I always, you know, target for decreasing risks associated with surgery, and then also improving outcomes afterwards. then post operatively, I usually see my folks at about 3 or 4 weeks. Kind of assess how they're doing. I always really want them to have like a high protein diet going home. As well. see where they get nutrition engaged outpatient if they need it."<br><br>Physician 5: "So I have an outpatient clinic for elective general surgery. I see patients on a weekly basis for new consultation in general surgery as well as my specialty area, which is minimally invasive. And for gut surgery, typically a patient would be referred to my clinic, usually by their PCP or gastroenterologists, though sometimes their self referrals of people that have found me online. They show up for clinic visit and we go through the initial consultation. I'll order any additional testing that needs to be done. If they're ready to go ahead with surgery, we go ahead and get them booked that day, otherwise I have them do their testing and then come back and see me for an additional pre-OP visit. They get scheduled for surgery through my scheduling team and then have their operation with me in one of the 3 locations that I perform. Surgery is either at the [BLINDED] Hospital or at the [BLINDED] surgery center depending on the type of surgery they have. They'll either have then a follow up visit. If it's a very simple small surgery. We typically do those follow ups by phone. And they're typically with my nurse practitioner. If it's for any larger surgery, they'll have an in person visit with me somewhere between 2 and 4 weeks after the surgery. Typically, with minimally invasive operations. Only one post out visit is needed. People tend to do pretty well. They don't have a lot of long term needs and |

| | | |
|---|---|---|
| | so they're then dismissed from the clinic with, Follow up Prn. If they have any new issues or concerns. Obviously, if I identify any problems in the post operative period, they're following longer with additional minutes. There's also a subset of patients that are not seen by me pre-operatively, the we have a robust nurse practitioner group that works with us. So for very simple things like umbilical hernias like palmers. They'll see our our feeding tubes. They'll just see our nurse practitioners in clinic. They'll do all of the initial evaluation, and then set them up for surgery with us, and then I meet them the morning of surgery. So there are a subset of patients that I only meet on the day of surgery, and I don't see it any other time." | |
| **Adherence to Clinical Regimen:** Capacity of patient to meet treatment plan and goals | Case Manager 1: "I would say the big ones tend to be like lack of support, whether that's if they don't have a lot of support people, or they don't have transportation or, you know, lack of resources. If you know their lower income or are not insured or under insured. Those are obviously big barriers and then additionally, more kind of generally on the medical side, would just be general compliance. which sometimes can just be an education gap, and other times is just patient preference will say, like, if they're just kind of not doing or following the intended care plan."<br><br>Physician 1: "I think there's, I mean, clearly, there's social determinants of health and kind of health literacy components social support, you know, and family members or loved ones who are able to encourage adherence to plans and treatment strategies. Certainly in the bariatric population there's a lot of metabolic component to their adherence to you know, durable weight management and diet patterns. Obviously, there's financial problems. But that's kind of encompassing what we talked about. But you know, I think travel can be challenging for our patient population, who sometimes is coming in from out of state just to see some of us. Yeah, those are the ones that come to mind." | Patient 23 (male, in their 40s, general/soft tissue, NH Black, lesser opportunity): "Yeah, she just told me I needed plastic surgery. Stuff like that is, her recommendation is to do the plan to switch it, because I've been coming to the hospital for like maybe 3 times they do surgery about the same armpit, and she says, you recommend me to do plastic surgery. And I say, Okay, I will talk to them, man. It's hard for me. You know what I mean. I'm working for Uber. You know. I'm driving. I don't know whether I will qualify to collect disable. You know what I mean? Because that's it's not a small surgery. It's a big surgery that would take like maybe 3 months to stay home, and I have family to feed. I got kids. I got mortgage to pay. So it's crazy. I have to. I have to plan that. It's not something I can just come do it. you know, doesn't work like that. And he referred me to the process. So you to talk with them. And I said, Okay, I will do that. And now I have a referral to go there to talk to them. So that's what we're saying. She's the one who will do the surgery, so she look at it and see that. Oh, yeah, look good. You know what I mean. He put the bandaid back again. I said, this |

| | | |
|---|---|---|
| | Case Manager 2: “Okay, challenges with adherence, I would say cost of medications. That's a big thing some of the patients, and particularly with our Medicare patient population. Some of the medications that they need are expensive. and are high tier on their plan think transportation is an issue for many of our patients. They don't live right here in [BLINDED] area. So that's definitely. A concern, a challenge. Yeah sometimes lack of local resources. It seems to be getting a little bit better. But you know a lot of times. The case managers, we primarily will set up home healthcare services and stuff for patients. And sometimes it's very challenging in certain areas to even find providers. So that can be an issue. Oh, and II think technology, too, for some of our for some of our older patients, I mean? You know I have a patient coming in today who's in his eighties, you know, having a gastrectomy and I don't, even though I haven't met this patient yet. But sometimes for some of our older patients, technology is an issue for them.” | is, look good. It's coming, you know. Start getting close. Sounds good.”<br><br>Patient 20 (male, in their 60s, gastrointestinal/benign, NH Black, most opportunity): “I did have some challenges as far as trying to coordinate after care. At 1st there, there was a lot of information coming at one time, and you have to decipher or all that information they're telling you. where you're gonna stay, what's gonna do with your medications. Here are some results. Here's what we want to do, and everyone had their own different process that they were doing. In particular. What I would like to see happen is that a continuity of care instead of just a Radiology does this or laboratory does this. I wanted to go across the spectrum. So laboratory does this. It ties into radiology this way. Then the next person gets the ball, and but unfortunately, some of the time it's not like, at least not at first. It wasn't like that until I kinda advocated for my own medical care. Then I see our time together.”<br><br>Patient 17 (female, in their 40s, general/hernia, NH Black, lesser opportunity): “I struggle with the weight loss a little bit, but I got through it. I actually didn't realize that I was at the mark, you know, that was a good thing. So yeah, it was just the weight part. That was the most struggle.” |

#### Supplemental Table 2. Current collection and management of health-related needs

| Theme | Care Team Quotes | Patient Quotes |
|---|---|---|
| **Collection of Social Need** | | |
| Awareness: Knowledge of health-related needs | Case Manager 1: "yes, I would say, all [PATIENTS REPORT HEALTH-REALTED NEEDS]. All, or most of the patients that I work with."<br><br>Advanced Practice Provider 1: "I'm not sure what that is the social determinant of health wheel. I'm not sure. I mean, I use the in basket all the time. And you know, for calls and messaging like my chart messages. We use that pretty much every single day. But I'm not sure what the the social wheel is."<br><br>Physician 5: "so they don't frequently offer it. No, it's not something that is routinely part of our assessment, really. And I always ask people the things that I do always ask, are are you gonna be able? Are you gonna have somebody that's gonna be able to help you out afterwards? But I don't go into the nitty, nitty gritty of the social situation. I've had that bite me a couple of times."<br><br>Resident Physician 1: "Yes and no. I mean, I think if we ask if we ask about it, and we ask about it because they're usually relevant to their condition. Yeah, I think they certainly do report it. You know, unless we're asking about it. I don't think it's like freely brought up often." | Patient 2 (female, in their 70s, gastrointestinal/benign, NH white, lesser opportunity): "I don't remember that. They asked. Well, they probably did. My gosh! Before? Yeah, before before I left. I don't think I would ask anything when I came in like that. And then, of course, my home nurse did ask me those questions, too. You know that I had but yes, I think everybody. I think I was asked those questions when I left both times down there was going to be okay at home. And you know, whatever."<br><br>Patient 6 (male, in their 60s, gastrointestinal/benign, NH white, most opportunity): "like I said. You know I was there a couple of 3 days. I said I need a shower, and they're very obliged, and they got me hooked up. We wouldn't, you know, have shower and everything. So I mean That's social needs, cause I was feeling pretty funny, and I, you know. But they hooked me up so."<br><br>Patient 8 (male, in their 30s, gastrointestinal/benign, NH white, lesser opportunity): "I don't think so. I don't know [IF SOCIAL NEEDS INFORMATION WAS COLLECTED] … Well, I guess I like I would have if it came up. I just can't remember, if it did yeah, it's nothing preventing me from talking about it… If I come to think of it, I mean, it might have come up with like, say, Case, |

| | | |
|---|---|---|
| | | manager. She might have asked about it.” |
| Roles: Care team member(s) involved in collection of health-related needs information | Case Manager 1: “Yes, as case managers outpatient. If we're calling patient for 30 days or more, we do a comprehensive assessment…..We assess you know, their medical and social needs, which includes I don't think all of those things but most of them, as far as you know housing, insurance, income.”<br><br>Social Worker 2: “So for all patients that like have any needs like homelessness, addiction, medicine like issues of transportation, etc. We'll get a consult from the nurse to do like our social work assessment, or I'll just do a chart review and kind of see that their needs and so I'll complete our social work assessment, just asking some questions about the social determinants of health and then giving. We have, like a really big resource packet. We give any specific resources for like housing and things like that.”<br><br>Case Manager 2: “We're more focused on the utilization review piece of it and all in the discharge planning piece. They're[Social Work) a little more focused psychosocially but there's a lot of overlap between us. But we are the ones who are initiating the discharge instructions for the patients communicating with all the members of the team we do. you know. Make sure they have the appropriate follow up appointments, scheduled and testing procedures. We do a lot of communication with pharmacies, with insurance companies so medication prior authorization is probably a big one for us. yes. So there's a lot of overlap. But our focus as case managers inpatient is you know the discharge plan and making sure that that we have a safe plan for discharge for the patients … We kind of do a basic, you know assessment. As far as you know, their living situation. How many stairs. They may have in their home. any transportation issues, you know. Do who do they live with? you know who is their support person, you | Patient 17 (female, in their 40s, general/hernia, NH Black, lesser opportunity): “Not by name. No, but I just remember it was a nurse. It was yes.”<br><br>Patient 4 (male, in their 70s, cardiovascular, NH white, lesser opportunity): “I believe. Oh, the case manager and the nurse! They both discussed it with me.”<br><br>Patient 20 (male, in their 60s, gastrointestinal/benign, NH Black, most opportunity): “Yeah, I spoke with a nurse. I spoke with case managers. When I say case managers, I spoke with case managers in patient advocate.”<br><br>Patient 1 (female, in their 70s, cardiovascular, NH Black, most opportunity): “I did talk to I can’t remember who it was but the care team…someone during the discharge is what that went over the information with me.” |

| | | |
|---|---|---|
| | know, if they live alone, can somebody stay with them for a few days after their surgery, if needed?”<br><br>Case Manager 4: “Well, we have an actual social determinant of health assessment that the social workers do. But part of my assessment is, you know, who do you live with? Is it a house or an apartment. How many stairs to get into the house. you know? Do you do those independently? Do you use equipment to use those? I mean? I have to. We paint a picture together? And I asked them. And I said, You know, I'm asking all these questions. I'm not. I'm just gathering in the beginning, cause, you know, maybe they've had a foot surgery in their bedrooms up stairs, so maybe I need to get them a bed for downstairs. Temporarily, things like that.” | |
| Documentation Process: Methods used to collect health-related needs information | Case Manager 1: “Both the inpatient and outpatient case managers have a comprehensive case management assessment that we use and there's a flow sheet that goes along with that.”<br><br>Physician 1: “I ask everyone some social questions. You know. To me what's most important is their smoking status and smoking history. And I asked everyone about their work or their overall day to day activity levels. I think that's important in a surgical practice. I sort of intentionally do not ask them a lot of other social things that could be that can be a little off putting to patients, I think, who are simply requesting a surgical procedure be performed. And a lot of times, I think. If they want to volunteer information. Talk about information I'm happy to. But, for example, I don't take a detailed sexual history or a detailed religious history or you know, maybe even just a relationship. History is not of interest to me, because I don't think I'm going to impact that for the patient dramatically. And if they have questions about that, that I'm happy to talk about it. But so I was asking about their work. And I say, what kind of work do you do? What kind of work did you do? And I feel that that's very important for | Patient 6 (male, in their 60s, gastrointestinal/benign, NH white, most opportunity): “I mean, I mean, we had small talk stuff. So you know, I mean, maybe they were asking me questions. I wasn't really realize that I was being asked. Okay, but they were like, you know, you know, we talk about the home environment stuff like that. So but I I I was taking it as small talk. But maybe you know, with the help it was more.”<br><br>Patient 21 (male, in their 50s, gastrointestinal/oncology, NH Black, most opportunity): “It was a series of questions that they would ask on the screen. There were 3 different people that come in and ask the same question one after the other.”<br><br>Patient 4 (male, in their 70s, cardiovascular, NH white, lesser opportunity): “Well, they we talked about it, and then I believe there's I've signed so many forms, then they asked some stuff about how our house, and |

| | | |
|---|---|---|
| | the patient relationship? you know, just in terms of me getting to know them and their family, and how I can help them best. And you know II regularly type in my note. This is a [in their 40s] avid golfer, you know, because to me, if I'm going to be fixing their abdominal core musculature, I need to think about the fact that this guy golfs every day. That's gonna impact the way I do surgery or what technique I use or what long-term outcome. I'm hoping for that patient which is different than somebody who's wheelchair bound, for example. And maybe it's, you know, retired whatever. So I always ask those questions."<br><br>Social Worker 2: "But yeah, I think I get a lot. I also do get a lot of information from my case manager, because, like I said. They do an assessment. They don't do like how I do a social distress. They just kind of do everything like. Where do you live like? Who's your family like met more the medical side. But don't let me know a lot of times about like healthcare power of attorneys that people need. That's a big thing. We try and do like patients. They come in. They're altered that mental status, and they can't make decisions for themselves, and they don't have healthcare attorney. Sometimes we'll have to go for a guardian to be able to make those decisions. But yeah. And now our assessments, the nurses kind of do a mini assessment of our assessment, the social determinants of health. And so that's how I get a consult. So if they could do that, like mini or assessment with the nurse, and if they trigger and transportation, housing, etc. Then I'll get a consult to do the assessment. So I would say, now it's more of the consult, or I also just do my own chart auditing like from when I was some pretty new to this I just graduated just my first job. But my preceptor did a really good job training me how to look in the chart for certain things. That might need a social work assessment. And sometimes I if I just feel like they needed or I get a consult and then they don't actually need anything. So sometimes I just do it, and they don't end | and I had to fill out some forms for that. You know, where we lived at. If it was 2 story, you know that kind of stuff."<br><br>Patient 20 (male, in their 60s, gastrointestinal/benign, NH Black, most opportunity): "No, they came to solve me, and I gave them the same information. But and they supposedly well, but not, there's no follow up…They just asked it and annotated it. And that was it." |

| | | |
|---|---|---|
| | up needing anything. But sometimes I'll just do it, knowing everything. It just depends on the situation."<br><br>Advanced Practice Provider 2: "There is the [ONCOLOGY] distress screen. Are you familiar with that? So that gets sent to all of the oncology patients? And then if they fill it out on my chart and it flags, it comes back and the flow is that you're supposed to call the patient. If there are flag responses and discuss referrals." | |
| **Management of Social Need** | | |
| Monitoring: how health-related needs information is tracked and assessed | Social Worker 2: "Okay. Yeah. Yeah. We were just talking about that. I had a social work meeting, or just talking about that [EPIC® SDoH Wheel]. I like. Oh, to be honest, I always kind of forget it's there. I just don't look at it as much. But I do. It is helpful now, I mean, like my coworkers kind of pointed out to me. I just always forget, but it is helpful when I do see it to see that there are needs that need to be met, that so I can move with the patient to assess any needs that they have."<br><br>Physician 2: "So I actually remember my patients, really. Well, so a couple so I don't like to document things that patients disclose to me that are sensitive that they wouldn't want to share. So sometimes I'll just put like a little plus sign in my in my history where I type, and that plus sign to me was like a Oh, yeah. Remember, I had that conversation. They were, you know, sexually assaulted by [parent in their childhood] or there, and so I kind of remember a lot of that occasionally. I'll if it's a very difficult situation where I know I'm not gonna remember, like a very complex or multi-layer story. I'll use the sticky tab like the one that only you can see when you open EPIC® doesn't show to other people. It just shows to you to leave a little note to myself….Well, those are for the things where patients are like, you know, don't feel comfortable sharing. But if there are other issues, then I'll send a message to like the | Patient 18 (female, in their 60s, general/hernia, NH Black, most opportunity): "Yes, we reporting it in new. Anything new and we feel we feel like something is not right. We have to notify the provider."<br><br>Patient 2 (female, in their 70s, gastrointestinal/benign, NH white, lesser opportunity): "Yeah, cause I thought, well, if I have any questions I'll just call down, you know, call the doctor, but but you know that's an inconvenience for them answering phone all the time, which I understand. But I have so many people."<br><br>Patient 5 (female, in their 30s, gastrointestinal/benign, Hispanic, lesser opportunity): "Yeah, I would at least call up either by calling them in or sending a message through like a portal." |

| | | |
|---|---|---|
| | PCRM to get some nutrition support or like. So it's like if depending on the situation. So if it's one of if it's a disclosure of, you know, violence or something like that where they're not comfortable sharing, especially if it's a hit or remote history. If it's something more recent, you know, I'll refer them to like the domestic violence, shelter something like that more readily. I've only had, I think."<br><br>Advanced Practice Provider 2: "I think it [Oncology Distress Survey] may end up in a flow sheet. That's a way to look at it, but it comes to our in basket. If you were the one who had ordered it.… Not a PDF, so maybe they are not very good with technical terms. Yeah, it'll have all what the question was, what their responses. And if it is a concerning response. There's like a red arrow next to it."<br><br>Case Manager 3: "well, within the chart, the EPIC® chart. We'll go and review, like what information's available from progress notes and things that the social workers and outpatient center, or whoever else. Because I've noticed lately that more providers are being involved in that and asking those questions. So I will review that I always try to review those kind of things when the patients are coming in, so that I know kind of what we're facing and what's been done." | |
| **Type of Social Need** | | |
| Basic: A non-medical need that affects their daily life, but does not require a comprehensive solution from a financial | Social Worker 2: "Yeah, I would say transportation to honestly a big one that I didn't, that we run into a lot. But the patients have rights home can't drive when they leave here. So I think transportation is a big one, and just making sure that patients have a safe ride home."<br><br>Physician 2: "I would say food security is the other big one for me, because of the needs for healing from surgery, you know, requires such a, you know. A good diet and high protein, and protein is expensive. You know, and a lot of the foods that people can | Patient 14 (female, in their 50s, gastrointestinal/benign, NH Black, lesser opportunity): "that would be excellent, because if I could have reported transportation problems, my son could have been at work today, and their bus could pick me up, brought me here took me up. That would have been perfect."<br><br>Patient 23 (male, in their 40s, general/soft tissue, NH Black, lesser opportunity): "Yeah Yeah, |

| | | | |
|---|---|---|---|
| | perspective. (E.g., food, transportation, medicine) | afford a little bit more like like pastas and rice and stuff don't provide that same kind of you know nutritional benefit.”<br><br>Advanced Practice Provider 1: “we do, I'm honestly not sure of, you know, if transportation is one of the official things that's collected, but it is something that comes up in our clinic a lot and, as I mentioned before. That a lot of our patients travel quite a distance. To get here, and transportation can a lot of times be a significant thing… Another way that we kind of get an idea about finances is because a lot of our patients get pre-OP medications and things like that. So there's a concern raised with those about how they're going to pay for those things. A lot. Some of the medications that we use are over the counter. So we might not necessarily ask that question specifically about finances, but it comes up when you talk about those things. So I think there are. You know, your standard like, you know.” | sometimes I need transportation to go, you know. Go home especially like if I I don't need it to come here to. You know I bring myself every time because I'm driving, but I need like, if I have a surgery, to go home, and I tell them they provide transportation for me to go home. Okay, you know they always did. They always, you know…Yeah, it [getting transportation assistance] happened. Yeah, it happened a lot. You know what I mean. My wife wasn't supposed to come and pick me up stuff like that. So sometimes I might do something, so I tell them I don't have a car. I need a ride, and they find a ride for me. They can call over, and they can pick me and take me home. So this is my, I think my 3rd time I do surgery. They always do that to me. They always help me with that.”<br><br>Patient 22 (female, in their 40s, gastrointestinal/benign, NH Black, lesser opportunity): “transportation especially for a lot of people that live out of town, because I don't live here.”<br><br>Patient 1 (female, in their 70s, cardiovascular, NH Black, most opportunity): “I guess the the food, I guess your you know the food that you're supposed to be eating afterwards. Also. I think. to the the prescriptions. Well, they do give you a a copy of the prescriptions. I I guess I wasn't clear on the types of medication I was supposed to take for cardiac.Know I have a transplant. I know those medications. but I wasn't sure of the new medications that I was supposed to be taking, for you know, for the for the surgery, for the heart I guess that would be one thing |

| | | |
|---|---|---|
|  |  | that, I guess, I would, I would like to know, of because if I came home I guess that they have. Let's see. What's how. How can I say that? I guess I would need to know what medications I was supposed to take for the cardiac part. Once I came home, the new medications I was going to be on. that that would be one thing I would want to know about. cause I wasn't clear on what I was to take, you know. Even when I went to the rehab center I wasn't sure which medications were the medications I was on for cardiac. Guess that would be one concern that I would have.” |
| Complex: A patient that needs extensive assistance with different factors of their daily life. They involve more comprehensive solutions from a financial perspective. (E.g., housing, finances, employment, education, social support, air quality, domestic violence) | Case Manager 4: “When I set up home, health care for a patient, one of the barriers is, they'll say is there a teachable caregiver in the home. If the patient doesn't live with anybody, or my kids live out of town. Blah! Blah blah! Sometimes the home cares will say, Well, then, we can't accept the patient, which well great. you know. So I try to work with a company that will teach the patient as well as I can.”<br><br>Social Worker 2: “The insurance thing is like, once you need like long-term care, you're gonna live at a facility you need medicaid and so a lot of times like I have one guy that's been here for a while, and we're trying to get him medicaid to be able to go to get the care he needs at a long-term facility. So that's a big one that I do a lot because the social workers do that with insurance issues or so that's the main one with insurance. And then, financially, yeah, we screen people. We have. I see someone that doesn't have insurance or like whatever we have our finance team screen them for Medicaid, and then we'll get that process started if they need it for discharge. Yeah. And some people yeah, like can't afford to pay the bills like, I said, we kind of just give excuse me the resources that we have that might be able to help them pay for that.” | Patient 15 (male, in their 40s, gastrointestinal/benign, NH Black, lesser opportunity): “Alright, to be honest. Like I told you several times. I never know that I’d want to get finance because I used to. I used to work to be in my biz, but after the surgery. but my schedule is 2 surgery. I gotta do first one, then a reverse. So it really really take a financial toll. Told them it did, because the first surgery, I never! I know it really take a toll on me because I, am not be able to go to work for months and months. And it's hard to get financial. It's hard to get the system down to anything they do.”<br><br>Patient 14 (female, in their 50s, gastrointestinal/benign, NH Black, lesser opportunity): “definitely they should ask about home safety. And is there someone that is gonna be there to help you with your medical needs and transportation cause. When you leave the hospital. You have a ton of appointments, and it's kind of stressful, because if |

| | | |
|---|---|---|
| | Physician 2: “So I think financial difficulties for patients is huge. You know, because they. You know, depending on what surgery I'm doing. They're going to need time off from work. And if it's a small analrectal procedure, it may just be one or 2 days, which may not have a huge consequence to most folks, but for some people who that like one day off, is the difference between being able to pay for dinner for that night, or something like you know that tight, you know. Those are very significant things that have to be, you know, thought about, and then all the up to my big abdominal surgeries, where they may need to be off for like 8 weeks, you know, 8 weeks off of work. Especially if they don't have access to things like short term disability or family medical leave something like that's huge. Right? That's like what 10% of their income for the year.”<br><br>Advanced Practice Provider 3: “Yeah, I think you know safety. And you know safety in the home. Both emotional and physical safety in the home and social support. Because we're in cancer care with family, you know, if they don't have family, do they have some other sort of emotional support? as they go through the process? I think those are kind of the most important.” | you can't drive, you have to have a family member to do it for you, and these appointments are back to back to back to back. And then you you know you're not. You're that you're off work, but your family isn't necessarily off work. So it can create a problem.”<br><br>Patient 2 (female, in their 70s, gastrointestinal/benign, NH white, lesser opportunity): “So I do have issues, and I and I do live alone. I didn't mean to. I don't know if you thought someone live with me, but I really don't have anyone that lives with me…Yeah [I had someone stay with me after surgery], just actually, the first night that I came home. Yeah, my cous, my cousin was here, but she had a dog, which the dog irritated me, so I said, I don't think I need you, I said I. My arms aren't broken and my legs worked fine, so I don't think I need anyone. I feel fine. I felt fine when I came home is really very strange for the surgery that I had. So I I didn't expect to feel so well afterwards. But yeah, I did.”<br><br>Patient 8 (male, in their 30s, gastrointestinal/benign, NH white, lesser opportunity): “Yeah, I was just gonna say, I, I think that like making sure the people are around and then also making sure the the financial side of things. Are okay for the patient.” |
| Resolution: Actions to address unmet needs which may involve the use of community resources | Case Manager 1: “I was just gonna say, everyone can see our notes so it depends. What their needs are. If it's something more clinical than that would be something the case managers would manage generally, unless it's something a little more simple. If it's you know a more social need, we would consult and provide handoff to the social workers. Specifically, if it's a medication | Patient 23 (male, in their 40s, general/soft tissue, NH Black, lesser opportunity): “Yeah. They asked me. They asked me if you need a ride…Yeah [before I left the hospital]. After the surgery they asked me whether you need a ride. If you do need a ride, let us know. Okay, we'll provide for you Always easy. I don't have no |

| | | |
|---|---|---|
| | need, I usually reach out to our medication assistance program for assistance there.”<br><br>Social Worker 2: “Yeah, so we feel like, it's kind of a false narrative. Some people think that we can like get them an apartment which, unfortunately we can't. I don't really have the time during the day. Unfortunately, so, if they're homeless, we have like a homeless hotline. We give them and then for the housing resources we have lists of like section 8, housing and apartments. That we give to the patient and kind of just say I'm not sure like what's available for these departments. But here's like kind of a starting point but it's kind of it, or like a lot of people have like eviction issues which we try to do as much. We can't really do much with that. Unfortunately, I just give them the resource packet and say, here's some organizations that might be able to help you. I try not make any promises, because some of these organizations unfortunately run out of money pretty quickly.”<br><br>Physician 3: “I don't know who I would refer somebody to.”<br><br>Case Manager 4: “So there's actually a relatively new process. We used to. Honestly. Just give em a piece of paper and have them start calling around. There's a new process where we can make computer referrals. It's called UNITE US. However, you can ask and it goes by zip code that you put in the need for, say food. And it goes for in their zip code, and it'll show you what's available in their Zip code. The issue may be there isn't availability close by. They might have to drive to that food bank. They might have to carry a bag of groceries or that service is so full it might have a 2 week waiting list if that makes sense.” | problem with that always easy. They provide for me. I get down, they take me home…Yeah, they just, you know, brings comfort, and I go home. That's the only thing they do. They distract me. If you might distract paper, you know. Walk me out out of the hospital, you know, kind of get in. They give me my thing, and they tell me bye-bye, see, you know, and I go home… Yeah, of course I would use it. If I did it, I would definitely use it. I don't need it. You know what I mean? I'm not those kind of guys. You know what I mean. So if I did need it, I don't have no car, I would call them and tell them, hey, I need a transportation to come in on my appointment stuff like that, so they will help you provide car for me, I will go to my appointments”<br><br>Patient 20 (male, in their 60s, gastrointestinal/benign, NH Black, most opportunity): “You know, I and I put this question out several times. It's not that we need it. But I wanted to get my wife reimbursed, so let me explain. There's a place, I guess, an agency, and I haven't looked at. It's called Ohio job family services, or something like that. And this agency supposedly reimburses individuals. Family members for the help. But my wife is doing a fantastic job, and I pay on myself. But it's the same money. So I said, let's see if you can get. So a friend of mine said, Hey, [DEIDENTIFIED SOCIAL WORKER] can get reimbursed for her time and effort. I'm like, really, okay. So they said, call them. I've asked that question 4 or 5 times through my care team through the Non care team. I've never got a response to that question of who should I go |

| | | |
|---|---|---|
| | | through? Is there a telephone number, and I know this is not the 1st time that the career team and the hospital has heard that you know how what agency reimburses for that I know some. I guess it's called Medicaid. Aand I know I don't qualify for that, you know, and and rightfully so, I mean, you know. Cause they said, It's $24,000. I thought it was a month that you make. And I like, Oh, yeah. And then they said, No, sir, it is for a year, I'm like, Oh, no, I make a little more than that. And so you know. But there needs to be something like that in place where somebody follows up. If you, if a person has a question. Don't just take down information, follow up either in my chart, which is a very expedient way, my chart, or send a person a message, or you have their numbers. Give them a call."<br><br>Patient 7 (male, in their 60s, gastrointestinal/oncology, NH white, lesser opportunity): "No, I just thought it [collecting health-related needs] was a base research of history and stuff like that." |
| **Perceived Impact of Managing Health-related Needs** | | |
| Benefits: Perceived positive effects from the collection and manageme nt of health- | Advanced Practice Provider 2: "Super important, because the knowing, you know that the patient's going home to a safe environment, and they have access to what they need is very important for their post up recovery for one, which is, you know, the primary concern for what they're here for right now, but then, also just for an overall health benefit having support or being referred to resources to get what they need | Patient 1 (female, in their 70s, cardiovascular, NH Black, most opportunity): "I think this is an excellent idea Cause, I think the feedback is good, and and I think it makes the patients feel like they're being heard. you know, as far as the needs that they need. So I think this is a excellent idea as far as communication." |

| | | |
|---|---|---|
| related needs | to take care of their help in their overall just day to day is important for patients."<br><br>Case Manager 4: "Honestly, it can make or break the success of the overall care because you can say: Hey, we're gonna do this as a doctor. We're gonna do this, this and this cause that. What that's what needs to be done. If they don't have food to heal that wound. It's not gonna heal right if they don't have clean water. if they don't have transportation or insurance information or somebody that can help them. Then they might have a setback like I said, in the perfect world everything I lined out for you on that piece of paper or that care plan is smooth. That never happens, you know that. But there's a ton of factors that we try to address even before they leave the building, so that if there is something that I've anticipated potentially could come up. this is the resource that you need to call for that make sense."<br><br>Case Manager 3: "Yeah, I think there's a real need for it, and I'd like to see a lot more screening done before the patient comes in. They're here for such a short time. And a lot of times we're just starting the process. We don't, you know especially these social things. I mean. It takes a while to get them processed through the system, whatever that is. You know whether that's meals on wheels. Making referrals to the [DEIDENTIFIED CANCER CLINIC] or to their whatever that that doesn't happen overnight, and most of the time we try to make it happen overnight. But but you know the social, the more social or economical issues, such as getting them hooked up with say, energy, assistance for housing, heating in their home, or even getting them help with : say, apartments or housing costs you know a lot of times they don't get that identified till they're here. And they're saying, Okay, what are you going to do about it? Well you know, that's hard, because they're here for a short post. Stop course. And we can. We can refer them out. But it's not gonna get. It's not going to get solved while they're here. But if there's | Patient 7 (male, in their 60s, gastrointestinal/oncology, NH white, lesser opportunity): "So inside, it's a necessity. you're not trying your best. If you're not doing that. Okay, you know you're shorting yourself. All you're doing is you're shorting yourself. You don't give them the full story."<br><br>Patient 1 (female, in their 70s, cardiovascular, NH Black, most opportunity): "the first 5 days when I came they said I'd need someone.They're with me 24/7, for you know, for a week. So they told me all that, and I had all that planned out, which was like. I said it was perfect, because they told me what to expect when I left the hospital. That helps me plan and go ahead." |

| | | |
|---|---|---|
| | any way we could start it beforehand. I think that we at least it would be in process, and they wouldn't have that stress of thinking about that during while they're recovering from surgery.”<br><br>Advanced Practice Provider 1: “I think it's very useful to kind of plan ahead, like I had mentioned. I think it's also useful to use our resources. Our case managers are kind of our, you know, right hand man, so to speak. We work very, very closely with them. they, you know, we'll pull from this information, have conversations with needs that we pick up and you know, it's it's one of those things where you follow also, too, because things can, you know, change. You can look back at that social history and kind of non medical things, and you know, at one point in time somebody may have you know family support. And eventually, maybe, that family support has since passed away, or something. So you can. There, it's it's dynamic, and you can always see you know those changes. But you have that initial base point. So it it is helpful.” | |
| Barriers: Perceived negative effects due to challenges in the collection and manageme nt of health-related needs | Case Manager 1: “Generally it's pretty similar a lot of times, you know, you can call and make the referral. Sometimes there's an online form to do it. I mean, it is variable. But in general it's the idea itself is fairly similar. But sometimes like, especially with some of the charitable pharmacies, it is a little more complex because they'll need an application and then like income information for the patient. So then they have that part that they have to do themselves cause obviously we don't have that information. The patient would have to send that …. Yeah. Yes, in multiple ways. You know, sometimes they struggle to actually just send it. You know, if they don't have or have access to like a scanner or something if it's just a fax they don't have access to. I have had patience like email it to me, and then I can fax it. Sometimes they'll bring it physically into us, and we can fax it for them that way. And then other times. Patients don't want to fax or send in their income information. And then that's tricky. I | Patient 19 (male, in their 30s, gastrointestinal/benign, Hispanic, lesser opportunity): “So, he was basically saying that like he didn't get like, why, it would be important to like share your social needs with the doctor, because he says like, even though he would have problems, he wouldn't specifically communicate it with doctor…He just wouldn't feel comfortable telling. It's out there like he needs this, or he needs that. He just doesn't feel good… He just feels like sharing those needs isn't something that he wants to do or he would like to do. He doesn't feel comfortable.”<br><br>Patient 23 (male, in their 40s, general/soft tissue, NH Black, lesser opportunity): “No, I never ask them something. You know what I mean. I'm a patient. I don't |

| | | |
|---|---|---|
| | would say like moderate challenges are fairly common and I may be like, you know couple like once a month. you know. once every other month I'll run into them more like really significant challenges. I hope it would be less frequent than that. Given what I'm trying to do. But I mean that that's like an estimate. I know it's not like an everyday, or, you know, every other week occurrence. But it does happen more than like …"<br><br>Physician 1: "I think it's a sensitive issue in the surgical setting, because I think it needs to be tailored to the situation and it needs to be relevant to their care. And so I think inundating patients with extensive surveys of sometimes very personal information, I think. may have detrimental impact to patients."<br><br>Social Worker 2: "So we have this new thing called UNITE US. And it's like an online referral system. You can make are make referral. So the patient just has to agree. Sometimes they agree or they don't. It just depends. Then we have them sign a consent form. It's yeah, like month or 2, and it's like pretty new. So I haven't used it a ton. Some people just don't really want it one. So I've had a couple of people. But yeah, we make a referral, and then but it's kind of con complicated, because like depending on what I make there for all, and how long the patients here for they it's a lot of it's going to be when they leave the hospital. Those these organizations will be following up with them. So we still, we. Still, we're still trying to get clarity from our management of like, who's supposed to follow this after they leave, because we don't follow any patients when they leave the hospital because, and I really don't know. I haven't had any interaction on so patients. It's just to see kind of how it works. Once they leave a lot. It's just kind of slow, and how quickly it is. And both patients are kind of in and out relatively quickly."<br><br>Social Worker 1: "I can see the benefit of the patient doing it [DOCUMENTING | bother people, you know, as soon as I walk in the hospital as well. If I stay in hospital I don't buy a lot of things. I did it for myself. You know what I mean. If I want to go to the bathroom, I, you know, get up and go do it, you know. Stuff like that, I mean, I don't bother them a lot, even all the nurses. you know what I mean easygoing somebody, you know."<br><br>Patient 20 (male, in their 60s, gastrointestinal/benign, NH Black, most opportunity): "I think a lot of the information goes in, and that's [IT] and it just stops."<br><br>Patient 7 (male, in their 60s, gastrointestinal/oncology, NH white, lesser opportunity): "You know, I've been asked a lot of the same questions over and over again. So I was wondering why it wasn't all documented in my charts, so everybody could see the whole story. My story, and as thorough as I find my charts to be. and I really like that program. I found it a little bit redundant, but at the same time I was very patient about answering all the questions when it was asking." |

| | |
|---|---|
| | HEALTH-RELATED NEEDS]. But then how truthful are they being? Because they don't wanna like be a bad score. do you know, I don't know what's a good question. I think there could. I think there's benefit to both….I think some people who are really struggling. This is very honest, and they're like, no, I'm struggling, and this is why so I think if it's really not as good, they're very honest. I think it's those people that may be in the middle that'd be like, well, so I really know everything's fine. Don't worry about it. We'll figure it out. So I think we experience more of that cause. I think I have both extreme where they're they're fine. They really don't need much. And then the ones that do need more. They're okay with sharing everything. I think it's more of those middle people that they don't want to ask. But they're not sure." |
